# Identifying medications underlying communication atypicalities in psychotic and affective disorders: A pharmacovigilance study within the FDA Adverse Event Reporting System

**DOI:** 10.1101/2022.09.05.22279609

**Authors:** Michele Fusaroli, Arndis Simonsen, Stephanie A. Borrie, Daniel M. Low, Alberto Parola, Emanuel Raschi, Elisabetta Poluzzi, Riccardo Fusaroli

**Author notes:** Corresponding author: Michele Fusaroli.

## Abstract

**Purpose:** Communication atypicalities are considered promising markers of a broad range of clinical conditions. However, little is known about the mechanisms and confounders underlying them. Medications might have a crucial, relatively unknown role both as potential confounders and offering an insight on the mechanisms at work. The integration of regulatory documents with disproportionality analyses provides a more comprehensive picture to account for in future investigations of communication-related markers. The aim of the current study was to identify a list of drugs potentially associated with communicative atypicalities within psychotic and affective disorders.

**Method:** We developed a query using the Medical Dictionary for Regulatory Activities (MedDRA) to search for communicative atypicalities within the FDA Adverse Event Reporting System (FAERS, updated June 2021). A Bonferroni corrected disproportionality analysis (Reporting Odds Ratio) was separately performed on spontaneous reports involving psychotic, affective, and non-neuropsychiatric disorders, to account for the confounding role of different underlying conditions. Drug adverse event associations not already reported in the SIDER database of labeled adverse drug reactions (unexpected) were subjected to further robustness analyses to account for expected biases.

**Results:** A list of 291 expected and 91 unexpected potential confounding medications was identified, including drugs that may irritate (inhalants) or desiccate (anticholinergics) the larynx, impair speech motor control (antipsychotics), induce nodules (acitretin) or necrosis (VEGFR-inhibitors) on vocal cords, sedatives and stimulants, neurotoxic agents (antiinfectives), and agents acting on neurotransmitter pathways (dopamine agonists).

**Conclusions:** We provide a list of medications to account for in future studies of communication-related markers in affective and psychotic disorders. The current test case illustrates rigorous procedures for digital phenotyping, and the methodological tools implemented for large scale disproportionality analyses can be considered a roadmap for investigations of communication-related markers in other clinical populations.

## Introduction

### The confounding role of medications on communication-related markers

Affective and psychotic disorders have long been associated with atypical communicative patterns - e.g., decreased emotional expression and flat prosody (Cummins et al., 2015; Parola et al., 2020). This awareness is widely used during the assessment of the disorders, and is increasingly investigated through automated voice and content analysis (Faurholt-Jepsen et al., 2018; Hansen et al., 2021; Low et al., 2020; Parola, Lin, et al., 2022; Parola, Simonsen, et al., 2022). The combination of new powerful forms of machine learning, pervasive smartphone data collection, and other sources of big data will allegedly identify historically elusive markers for affective and psychotic disorders and therefore enable more reliable diagnoses, continuous evaluation of symptoms, and perhaps even personalized treatment (Arevian et al., 2020; Ben-Zeev et al., 2019; Cohen, Cox, et al., 2020; Cohen, Schwartz, et al., 2020; Insel, 2017). However, communication is a complex phenomenon and its relation to specific disorders is not straightforward, with many potential confounders and ethical considerations (Albuquerque et al., 2021; Corona Hernández et al., 2023; Parola, Lin, et al., 2022; Rybner et al., 2022).

Medications, which are disproportionately associated with neuropsychiatric diagnoses and their co-morbidities, can affect not only mental health but also the communicative patterns in the patient. For example, commonly used medications with anticholinergic effects (e.g., antihistamines and antidepressants) can cause reduced salivation flow (xerostomia) and sedation of the mouth, which could cause dysphonia and difficulty in articulation (Haft et al., 2015). Another example: high D2R occupancy antipsychotics are administered to patients with psychotic disorders and are also associated with slower speech and increased pauses (de Boer et al., 2020). Therefore, it is often not clear whether the communicative atypicalities identified as behavioral markers of affective and psychotic disorders could be partially confounded by medications. Unfortunately, more general investigations of the associations between communicative atypicalities and medications are still sparse, and no comprehensive overview is available (see Supplementary Material 1 – Section A for an overview of studies assessing the effect of medication on speech patterns in schizophrenia).

Therefore, the objective of the current study was to identify a list of drugs that could be associated with communicative atypicalities, which should be evaluated in the future as potential confounders in communication-related markers of affective and psychotic disorders. After introducing our two key sources of information– clinical-trial-based information (SIDER, Kuhn et al., 2016) and spontaneous reports (FAERS, FDA, 2022)–, four common causal mechanisms underlying observed associations between drugs and adverse events are briefly discussed. We present how the potential biases highlighted can be accounted for in the analyses before detailing materials and methods. Finally, the resulting list of drugs associated with communicative atypicalities are reported and discussed.

### Information sources

As medications are tested in clinical trials, adverse drug reactions are evaluated, and if the drug is approved for market distribution (marketing authorization) these adverse reactions are reported by law in the insert of the medication package (Poluzzi et al., 2012), also known as prescribing information or summary of product characteristics. However, as the drug is used outside of clinical trials (post-marketing phase) unexpected adverse drug reactions are often detected. For example, an adverse drug reaction could arise in populations not investigated in clinical trials (e.g., older or younger cohorts, pregnant women, patients with additional comorbidities). In addition, multiple drugs are often administered together (polytherapy), and an adverse drug reaction could arise from their interaction. Such suspected adverse reactions to drugs can be spontaneously reported to the regulatory agencies by physicians, marketing authorization holders, and the general public. Disproportionality analyses are statistical techniques developed to detect patterns within spontaneous reporting systems’ databases in an attempt to provide a more comprehensive safety profile of medications (Alves et al., 2013).

Clinical trials and disproportionality analyses have complementary strengths. Clinical trials have obvious advantages, primarily that, by carefully selecting homogeneous samples and randomly distributing them across interventions, they remove many possible confounders and provide a strong causal assessment. Conversely, spontaneous reports can cover a much broader variety of patients and drug uses, including adverse reactions that are commonly underreported during clinical trials, although certain causality cannot be inferred due to confounders and lack of randomization. For instance, rashes are easy to observe, and arrhythmias could be fatal. Therefore, both are relatively prominent in clinical trial reports (Loke & Derry, 2001; Seruga et al., 2016) as compared with symptoms such as raspy vocal quality, or mispronunciations of speech sounds. However, communication impairments can be disabling from the patient’s point of view, and therefore be more likely to be spontaneously reported, as has been shown for stuttering (Ekhart et al., 2021; Inácio et al., 2017; Toki & Ono, 2018; Trenque et al., 2021).

Pharmacovigilance has long acknowledged that spontaneous reports provide very noisy information riddled with well-known biases. For example, reports may be incomplete or duplicated, lack quality control of the information provided (e.g., patients do not have the right language and knowledge to accurately describe their symptoms), contain potential biases, and may ignore external factors such as the novelty of a drug and how media coverage of adverse reactions affects the number of reports (Poluzzi et al., 2012; Raschi et al., 2018; Wisniewski et al., 2016). In other words, causal connections between drugs and adverse reactions should not be established based solely on spontaneous reports. Nevertheless, by taking these biases into account, disproportionality analyses can generate hypotheses for further investigation in analytical studies (cohort and case-control studies). Finally, with large enough sample sizes, there are methods for approximately estimating the causal effect of drugs in observational studies by adjusting for these newly considered confounders through confounding-adjustment methods (Hernán, 2018). It should be noted that package inserts and spontaneous reports do not exhaust the possible sources of information on adverse drug reactions, which would include, for instance, the scientific literature, health records, and clinical expertise in general.

### Causal models underlying drug adverse event associations

Disproportionality analyses identify adverse events that are more frequently present in reports about a given drug than in reports not containing that drug. However, the observed association could be generated through different causal mechanisms, with four common ones represented in **Figure 1**.

**Figure 1.**
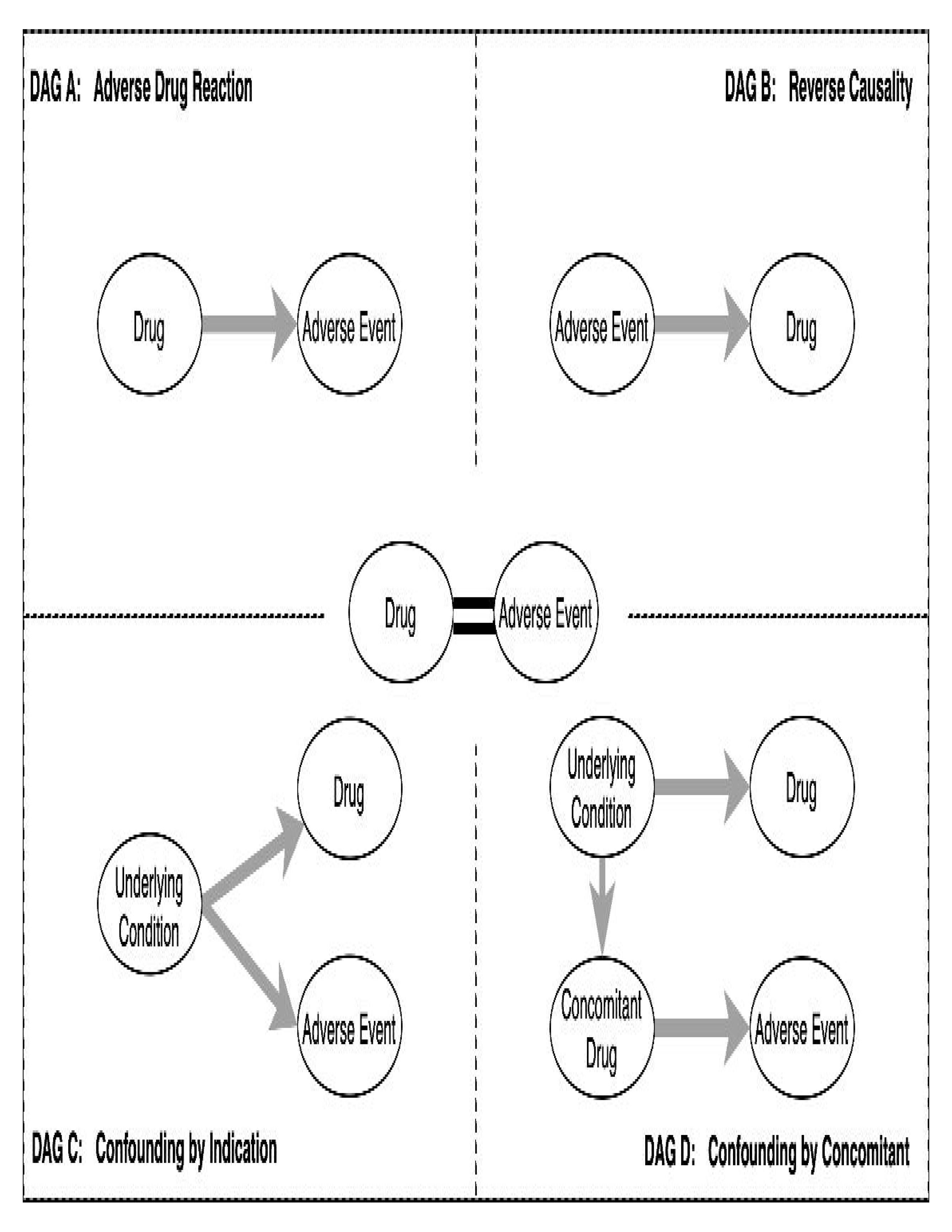
common causal mechanisms underlying drug adverse event associations. The four diagrams (A-D) represent four possible mechanisms which can all give rise to the observed association (in the center). The diagrams are direct acyclic graphs (DAGs), that is, graphs in which the nodes (ellipses) are the observable phenomena, and the arrows are the causal connections (which can only be acyclical, that is, go one direction and not form loops). **DAG A** represents the case in which the event is an actual Adverse Reaction caused by the administration of the drug of interest. **DAG B** represents a case of Reverse Causality, in which the drug is administered to treat the adverse event but is incorrectly reported. **DAG C** represents a case of Confounding by Indication, in which the underlying condition that justifies the use of the drug also more frequently induces the adverse event. **DAG D** represents a case of Confounding by Concomitant, in which the adverse event is a reaction to a co-administered drug (administered for the same condition or a related comorbidity).

The first possible causal model is simply that the adverse event is indeed caused by the drug (an **Adverse Reaction** to it; DAG A). For example, administering anticholinergic drugs often results in reduced salivation flow (xerostomia) and sedation of the mouth, which can cause speech impairment (Haft et al., 2015).

However, the association might also result from **Reverse Causality** (DAG B): the drug is taken because of the event (e.g., to treat it)^1^. For example, botulinum toxin is approved to treat spasmodic dysphonia, and antipsychotics are administered off-label to reduce stuttering (Maguire et al., 2020). These drugs can be reported as associated with a speech impairment because, for example, the lack of specific fields for symptoms of the underlying condition or for comorbidities often generates ambiguity in the reported information. Furthermore, when therapy does not reduce symptoms, reports might incorrectly record the indication for use (pre-existing stuttering) as an adverse reaction (after drug administration, stuttering is still there).

A third common possibility is the so-called “common cause” or fork (Pearl, 2009). Here the underlying condition is causing both the prescription of the drug and the adverse event, without there being any direct causation between the latter two (**Confounding by Indication**; DAG C). For example, psychotic disorders can involve some degree of communication impairment (e.g., alogia, i.e., reduced and vague speech, or disorganized speech), as well as the administration of antipsychotics. Therefore, when assessing all reports on FAERS, one might find an association between communication impairments and antipsychotics simply due to their co-presence, even if there were no direct causal association. Another example of the “common cause” problem is seen with gastroesophageal reflux, for which proton pump inhibitors (PPI) are administered. Acid reflux can also affect the larynx and vocal cords, resulting in dysphonia (Lechien et al., 2017), which would then appear to be associated with PPI even in the absence of a direct causal link.

A fourth common possibility is that the adverse event is indeed an adverse reaction, but to a different concomitant drug also prescribed due to the underlying condition (**Confounding by Concomitant**; DAG D). For example, diuretics are usually administered in conjunction with angiotensin-converting enzyme inhibitors (ACEI), which are known to cause bradykinin-related cough and laryngeal irritation. Therefore, diuretics might appear to be associated with dysphonia, even if the latter were exclusively due to ACEIs.

Finally, the relationship between a drug and an event may also not be reducible to one DAG only. Botulinum toxin may indeed be used to treat spasmodic dysphonia (DAG B), but it was also subject to a warning by the FDA because the systemic spread of the toxin can lead to temporary flaccid paralysis and related dysphonia (DAG A) (Kuehn, 2009).

### From causal models to statistical analyses

When disproportionality analyses identify an association between a drug and an adverse event, how can one discriminate between the possible causal mechanisms? It turns out that there is no replacement for clinical and scientific knowledge, including evidence from previous studies, clinical expertise, and informed mechanistic hypotheses. This knowledge must play a meta-statistical role in guiding the construction of statistical analyses. In other words, it is up to clinically and scientifically informed disproportionality analyses, not statistics alone, to identify plausible directions of causality and the necessary follow-up studies.

Specifically, reverse causality (DAG B in Figure 1) could be anticipated by carefully considering which drugs are used to treat the condition investigated. For instance, one could run analyses only on reports that do not include drugs used to treat communication disorders. Similarly, clinical expertise can identify whether underlying conditions are also likely to cause the adverse events of interest (Confounding by Indication, DAG C in Figure 1). This is the case of psychotic and affective disorders being associated with communicative impairments (e.g., flat prosody for both types of disorders, and semantic incoherence for psychotic disorders). A solution to this bias is to explicitly include the common cause in the model (“blocking the backdoor path” (Pearl, 2009)), for instance, by analyzing the populations separately: in our case, this implies separately analyzing individuals with affective disorders, individuals with psychotic disorders, and individuals without any neurologic medication in order to test whether patients with, e.g., affective disorders on vs. off a specific drug display higher rates of the adverse event of interest. By looking at reports for individuals not taking any neurologic medication, it is possible to exclude (and therefore correct for) psychiatric patients as well as other communication-impairing conditions such as anxiety, Parkinson’s disease, and dementia. This analysis is, of course, a first approximation: affective and psychotic disorders are complex conditions with very heterogeneous clinical profiles, comorbidities, and therapies. To move one step further, one could identify other underlying comorbid conditions likely to cause communicative impairments and produce a control analysis where all these conditions are excluded. Similarly, in the Confounding by Concomitant case (DAG D in Figure 1), one could identify drugs known to produce communicative impairments and exclude reports containing these drugs from the analysis. This also deals with what is known as “competition bias” (Raschi et al., 2018): known adverse drug reactions are easier to detect and therefore reported more frequently. Thus, established adverse drug reactions result in stronger associations, which mask the less reported unexpected ones. When known signals are removed, new associations may become visible.

While these techniques provide information about potential mechanisms, they do not guarantee accurate causal inference. Nevertheless, they contribute to the collective construction of more accurate knowledge on the relationship between drugs and communicative impairments by providing hypotheses to be explored and assessed in future investigations.

## Methods

### Overview of the analyses

The general pipeline of the analysis is represented in **Figure 2**, the details of which are explained in the following paragraphs.

**Figure 2.**
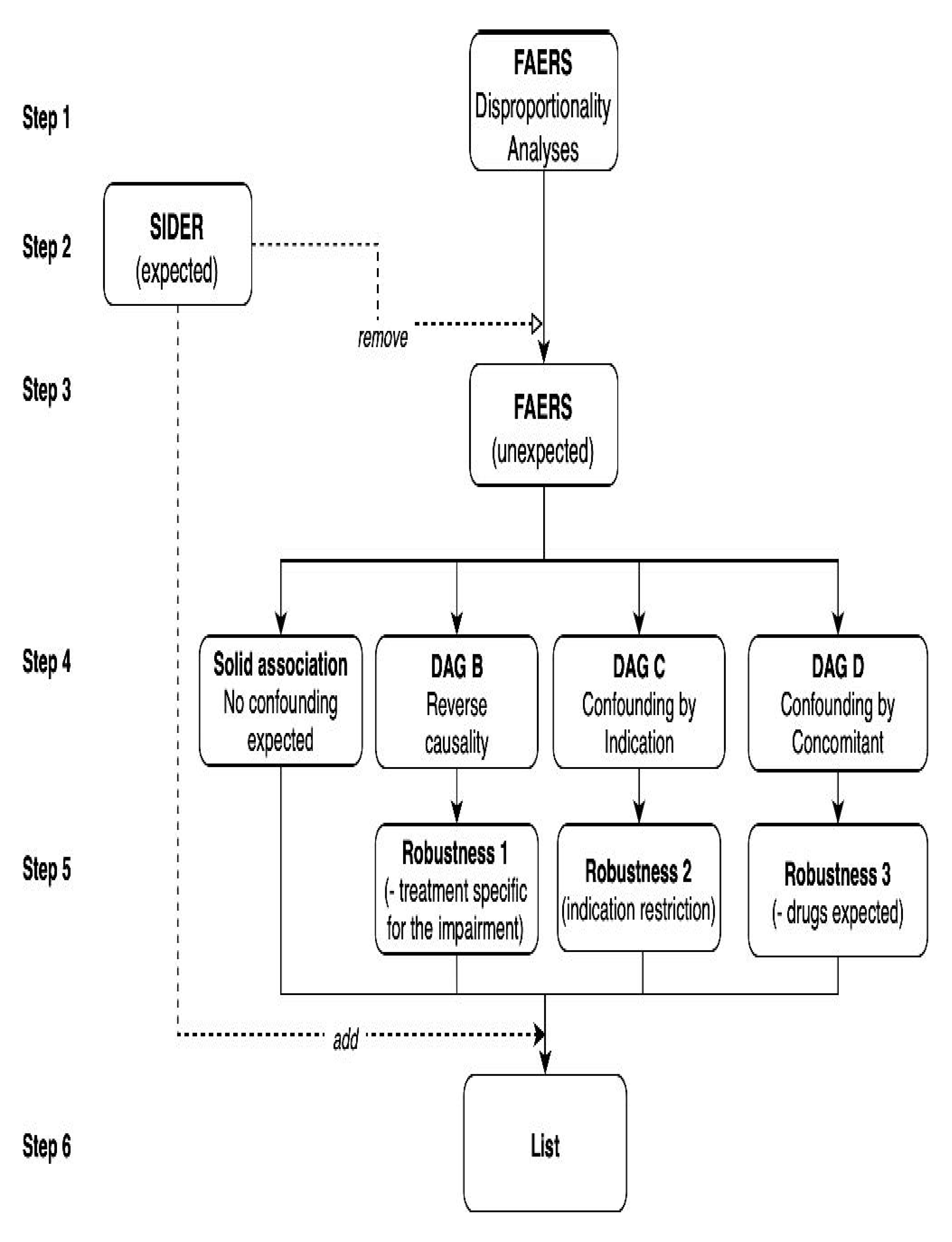
Analysis pipeline. Each step of the analysis is represented as a block and arrows indicate the flow of data from one step to the other. Descriptions of each step are provided in text.

### Definition of search terms

We relied on two information sources: SIDER for clinical trial reports of adverse drug reactions and FAERS for spontaneous post-marketing reports. Both sources employ a standardized hierarchical lexicon to code for adverse events, the Medical Dictionary for Regulatory Activities or MedDRA® (an international medical terminology developed under the auspices of the International Council for Harmonisation of Technical Requirements for Pharmaceuticals for Human Use – ICH). In MedDRA, the highest organization level is the System Organ Class (SOC, e.g., nervous system vs. vascular disorders), followed by the High-Level Group Terms (HLGTs, e.g., neuromuscular vs. neurological disorders), followed by High-Level Terms (HLTs, e.g., muscle tone abnormal vs. motor neuron disease) and Preferred Terms (PTs, e.g., hypertonia vs. hypotonia). Both SIDER and FAERS code their adverse events as preferred terms.

The MedDRA lexicon has some limitations that are important to acknowledge. First, the MedDRA does not always include the most adequate terms to report a given adverse event; therefore, some events are less likely to be reported or are reported relying on only partially relevant terms. Second, the same event may be reported using different MedDRA terms, often coded in different branches of the MedDRA dictionary. For instance, speech sound disorder is coded as a psychiatric event among “communication disorders and disturbances”, while dysphonia is coded as a respiratory event among “respiratory, thoracic and mediastinal disorders”. Third, and even more concerning is the limitation ensuing from the inconsistency between the MedDRA and different relevant conceptual frameworks to understand communicative impairments. The current study must rely on the MedDRA lexicon, since both its information sources - SIDER and FAERS - code their adverse events as MedDRA preferred terms. However, caution is needed in interpreting MedDRA terms in this study. The MedDRA has been developed to facilitate the identification of signs and symptoms - emerging as drug related adverse reactions – by a broad range of users with a diverse set of expertise: from clinical practitioners to patients and caregivers. Therefore, its terms do not easily map onto other very relevant frameworks, such as the categories of communication impairments as investigated within psychotic and affective disorders, and the nosological entities of the speech and language pathology community, built to systematize knowledge on commonly co-occurring signs and symptoms and their underlying mechanisms. For instance, while impairments in prosody are commonly considered a speech motor control issue in the domain of speech and language pathology, in the study of affective and psychotic disorders it is associated with flat and blunted affect, that is, with emotional aspects of the conditions. This implies differences in how prosody-related impairments would be categorized.

In order to partially overcome these limitations and to tailor the categories to the question at hand, good pharmacovigilance practices rely on the so-called Standardized MedDRA Queries (SMQs), which are expert-validated search queries that aggregate many partially overlapping preferred terms across the MedDRA to identify and retrieve cases of interest. In the absence of an SMQ for communicative impairments, six clinical and domain experts (pharmacovigilance experts, speech-language pathologists, psychologists, and experts on voice markers of affective and psychotic disorders; see the list of co-authors) independently clustered the communicative PTs based on semantic overlapping, and disagreements were discussed among the team until resolved. The multidisciplinary team developed the clusters considering that diverse and not necessarily expert reporters may use multiple terms to identify the same communicative impairment. Additionally, we excluded from disproportionality analysis generic PTs (e.g., speech disorder), which often imply low specificity in the report and could in principle indicate any communicative impairment.

### The FDA Adverse Event Reporting System (FAERS)

The United States Food and Drug Administration (FDA) Adverse Event Reporting System (FAERS) collects worldwide spontaneous reports of suspected adverse drug reactions and offers the highest accessibility to the public for customized analyses. Specifically, its raw quarterly data include demographic, therapeutic, and outcome details for each individual report. The reaction (adverse event) and indication (why the drug was administered) fields are standardized using MedDRA preferred terms.

The entire FAERS - Quarterly Data (FDA, 2022) (January 2004 to June 2021) was downloaded and pre-processed to remove duplicate reports and standardize PTs and drug names. For the standardization of PTs, we used MedDRA 24.0. For the standardization of drug names, we used the WHO drug dictionary accessed in March 2020 and iteratively integrated to include newly marketed active ingredients. Furthermore, drug names were linked to their specific code from the Anatomical Therapeutic Chemical (ATC) hierarchical classification (2022 version) to allow group visualization of similar drugs. Because individual substances can have multiple codes related to distinct indications of use or administration routes, we selected only one code for each active ingredient using a semi-automatic prioritizing algorithm (Gaimari et al., 2022).

### Exposure of interest

In order to identify medications associated with communicative impairments and deal with possible “common cause” biases (DAG C in Figure 1), we separately investigated three clinical populations: patients with a) affective, b) psychotic conditions, and c) without any neurologic medications (i.e., likely without any neuropsychiatric conditions, hereafter termed non-neuropsychiatric reports). To identify patients with psychotic and affective conditions we selected all reports that recorded, as a reason for using drugs, any PT (for example, ‘schizophrenia’) belonging to the HLGTs for psychotic disorders (“schizophrenia and other psychotic disorders”) and affective disorders (“manic and bipolar mood disorders and disturbances” and “depressed mood disorders and disturbances”). To identify patients without neuropsychiatric conditions, we selected all reports that did not include any neurologic drug (according to the ATC) nor any psychotic or affective preferred term. The results of the selection procedure are displayed in the Supplementary Material 1 – Section C – Figure S1.

Descriptive analyses were performed to characterize cases (displaying communicative impairments) vs. non-cases separately in the three populations of interest, with a particular focus on demographics, concomitants, co-reported events, and comorbidities (Supplementary Material 1 Section C). Differences between cases and non-cases may point to susceptibilities and potential biases not a priori acknowledged. For example, if we find that older people are more represented in cases than non-cases, this may point to a potential bias related to a higher frequency of speech disorders in the elderly.

### Disproportionality analyses for drug-event association detection

Disproportionality analysis (the analysis of a reliably more frequent reporting of an adverse event in presence of a drug than in presence of any other drug, Figure 2 – Step 1) was performed following good signal detection guidelines (Wisniewski et al., 2016). Using a contingency table 2×2, we calculated the Reporting Odds Ratio (ROR) whenever at least 10 cases of the event investigated co-occurred with the drug investigated. In fact, when few cases have been collected, the probability of detecting spurious associations is high. Considering a threshold of 10 cases allowed us to reduce this risk at the cost of missing some true associations for which not enough cases had yet been collected (e.g., for particularly novel drugs), and can be seen as an alternative to other conservative methods, such as the information component (Norén et al., 2013). The ROR was deemed significant when the lower limit of its 95% confidence interval was greater than 1. In other words, we report a potential association when the adverse event is more likely to be reported together with the drug of interest than with any other drug but the one analyzed.

We performed a disproportionality analysis evaluating associations between drugs (from the ATC 2022 classification, excluding mineral supplements and drugs included in the ‘Various’ class) and communication-related adverse events (sub-clusters of overlapping terms as identified in **Table 1**). The analyses were run on all reports involving a) affective and b) psychotic disorders, and c) non-neuropsychiatric reports. To filter out likely spurious associations, results were subjected to Bonferroni correction.

**Table 1.**
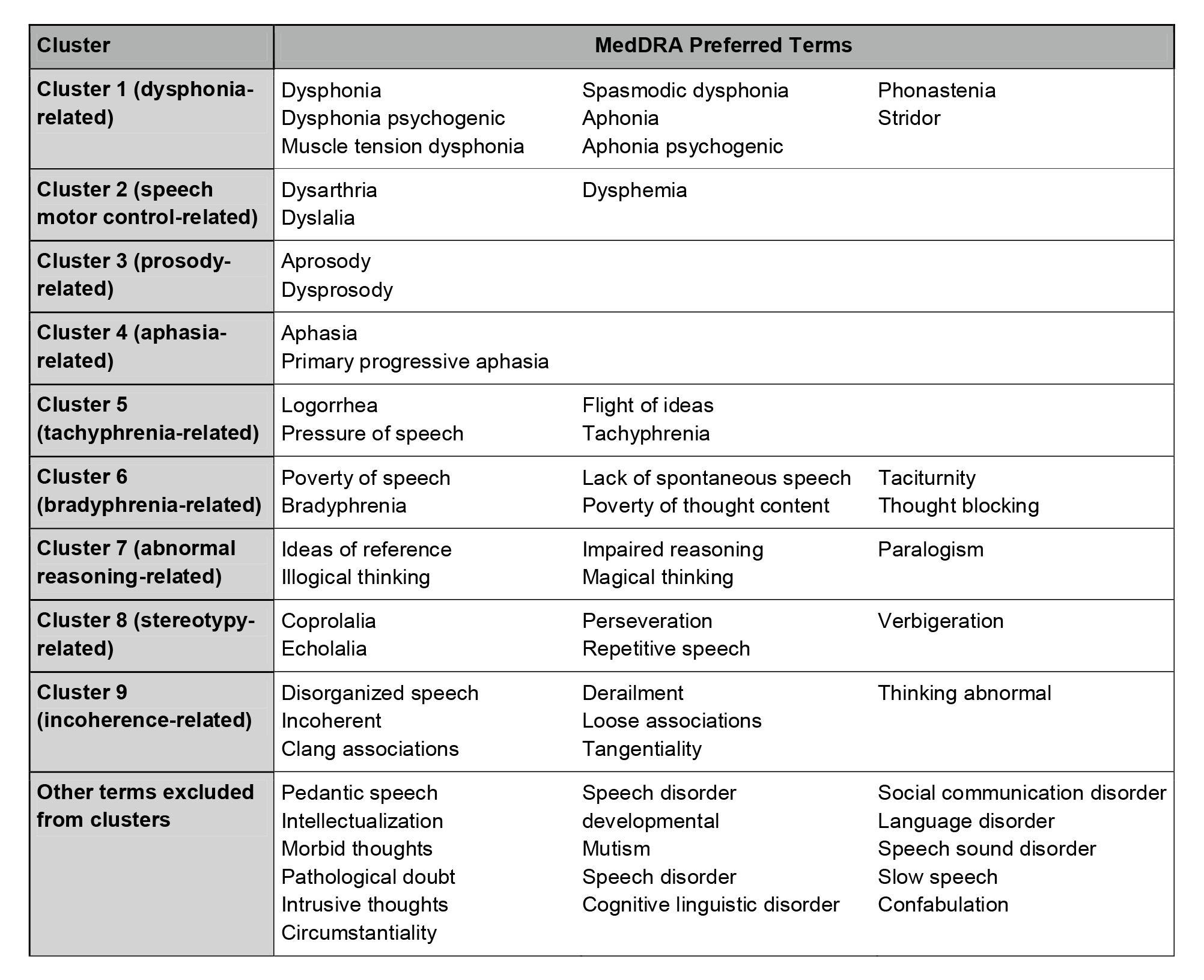
– MedDRA Query for the retrieval of communicative atypicalities reports. We identified multiple sub-queries including semantically overlapping MedDRA Preferred Terms. The clusters were obtained on the basis of semantic overlapping, and therefore on the possibility that the reporters may have used interchangeably different terms, rather than with a reference to existing speech and language pathologies.

### The Side Effect Resource (SIDER)

The Side Effect Resource is a public database that grants free access to the information contained in the package inserts, that is, the official information on a drug and its uses, in particular its side effects, compiled and distributed by the drug manufacturers. Package inserts are text-mined, and the information retrieved is coded using the ATC classification for medications and the MedDRA classification for adverse events.

For each subquery of potential adverse reactions, we searched the specific preferred terms. We considered the identified medications as expected associations (Figure 2 – Step 2), which did not require further discussion of potential biases and causal mechanisms. The associations found in FAERS but not present in the SIDER were considered unexpected and were further assessed for potential causal confounding (Figure 2 – Step 3).

### Robustness Analyses

Drugs unexpectedly associated with the sub-queries investigated were stratified according to expected biases (Figure 2 – Step 4) through clinical reasoning and according to the causal inference framework discussed in the introduction (paragraphs 1.3 and 1.4). We accordingly separated the associations into uncontroversial ones (plausible adverse reactions, Figure 1, DAG A, for which no specific confounder was expected), potential reverse causality (Figure 1, DAG B), potential confounding by indication (Figure 1, DAG C), and by concomitant (Figure 1, DAG D). Robustness analyses adjusted the estimates for possible confounders (Figure 2 – Step 5): excluding reports with the communicative impairment among indications or restricting the investigation to a specific indication, to account for reverse causality bias (DAG B; Robustness Analysis 1); excluding reports with pathologies that may be responsible for indication bias (DAG C; Robustness Analysis 2), at least for drugs that are approved for multiple indications; excluding reports with the drug responsible for the ambiguity to account for the concomitant bias (DAG D; Robustness Analysis 3). The procedure applied is documented in Supplementary Material 1 – Section D.

### Aggregating Results

The expected adverse drug reactions from the SIDER and robust unexpected associations from the FAERS were aggregated in a list per each main cluster of overlapping PTs, Figure 2 – Step 6. To provide a detailed overview of the results, we visualized each list as a table showing expected and previously unexpected adverse reactions organized according to the ATC hierarchical classification (see Supplementary Material 1 – Section D).

To provide a more general overview of the drug classes that should be considered for the analysis of communication-related markers, we built a heat map showing the associations at the third level of the ATC classification (e.g., antipsychotics, antihistamines, and antidepressants; see the public repository (M. Fusaroli & Fusaroli, 2021) for a collated heatmap at the level of single active ingredients).

## Results

### MedDRA query for case retrieval

We defined nine clusters of overlapping MedDRA PTs referring to communicative impairments. For simplicity, we named the clusters with reference to the semantic overlap and specific concerns (communicative impairments in affective and psychotic disorders) that guided the aggregation: related to 1) dysphonia, 2) speech motor control disorders, 3) prosody, 4) aphasia, 5) tachyphrenia, 6) bradyphrenia, 7) abnormal reasoning, 8) stereotypy, and 9) incoherence (seeTable 1). Sixteen of the communicative PTs were excluded from clustering and the subsequent analyses. It is important to note that the clusters might not be entirely coherent with current uses of the terms in the speech and language pathology community, but they were the result of an interdisciplinary consensus, and they will be discussed and clarified where they could generate misunderstandings.

### Populations of interest

We selected three populations of interest: 302,000 reports involving affective disorders, 11,631 psychotic disorders, and 7,703,183 non-neuropsychiatric disorders. A detailed presentation of the number of cases (reports with communication-related adverse events) and non-cases is presented in the Supplementary Material 1 – Section C Figures S1-S5 and Table S2.

### Expected and Unexpected Solid Associations

Disproportionality results and effect size for each cluster and each population are reported in the Supplementary Material 2. We detected both expected – according to the SIDER - and unexpected drug associations and performed robustness analyses on the latter ones. The result was a list of 291 expected and 91 unexpected potential confounding medications. Emerging results are shown in Supplementary Material 1 - Section C (Tables S3-S9, Figure S6). No association was found for the prosody and abnormal reasoning clusters.

We detected 72 drug classes (ATC third level) associated with a disproportional reporting of the dysphonia cluster: 53 were already expected based on the SIDER, 10 classes included both drugs already reported in the SIDER and unexpected drugs (integrated classes) and 9 were entirely unexpected. Restricting to strong signals (i.e., disproportions significant after the Bonferroni correction) in the non-neuropsychiatric population, the highest number of cases involved inhalants – fluticasone (4669 cases, ROR = 10.48, 95% CIs = [10.14-10.81]), salmeterol (3099, 12.16 [11.71-12.63]), and salbutamol (2434, 5.52 [5.29-5.76]), while the highest lower limits of the 95%CI of the ROR concerned two VEGFR-inhibitors –regorafenib (530, 22.25 [20.29-24.35]) and axitinib (437, 14.27 [12.92-15.37]) and salmeterol. The many anticholinergic drugs already present in the SIDER were integrated with unexpected signals for umeclidinium (an inhaled bronchodilators), rupatadine and fexofenadine (antihistamines). Among the robustness analyses implemented, we accounted for reverse causality (DAG B: botulinum toxin excluding its use for spasmodic dysphonia(Faham et al., 2021)), confounding by indications (DAG C: antihistamines restricted to urticaria, to exclude the confound due to asthma) and concomitants (DAG D: cardiovascular agents excluding angiotensin-converting enzyme inhibitors; beta agonists excluding inhalants).

We detected 37 drug classes associated with a disproportional reporting of the speech motor control cluster (17 expected, 10 integrated, 10 unexpected). The most numerous cases concerned immunomodulators used in multiple sclerosis – natalizumab (770, 4.48 [4.16-4.82]) and interferon beta-1a (674, 3.62 [3.34-3.91]) – and a selective calcium channel blocker – amlodipine (376, 2.05 [1.84-2.27]). Drugs with the highest lower limit were anti-infectives: vidarabine (20, 71.42 [42.66- 113.63]), valaciclovir (334, 12.14 [10.84-13.55]) and metronidazole (309, 9.48 [8.44-10.63]).

A total of 51 drug classes were associated with the aphasia cluster (19 expected, 10 integrated, 22 unexpected), with the most numerous being natalizumab (872, 5.54 [5.16-5.94]), interferon beta-1a (643, 3.71 [3.42-4.02]), and levothyroxine (327, 1.57 [1.4-1.75], and the highest disproportionalities being with antineoplastic such as CAR-T engineered cells used to treat hematologic neoplasia –axicabtagene ciloleucel (114, 43.68 [35.78-52.8]) and tisagenleceleucel-t (58, 24.5 [18.52-31.84])–and avapritinib (20, 18 [10.33-26.44]).

Concerning the stereotypy cluster, we did not find any unexpected signal and only four expected drug classes: antineoplastic (ifosfamide), antiepileptic (topiramate), antiepileptic (phenelzine and bupropion), and contrast agents (iopamidol). The only strong signal was with interferon beta-1a in non-neuropsychiatric patients (20, 6.12 [3.65-9.73]).

A total of 12 drug classes were associated with the tachyphrenia cluster (4 unexpected, 2 integrated, 6 expected). We observed associations based on only few cases, the greatest being clarithromycin (49, 22.38 [16.43-29.8]), levothyroxine (47, 2.29 [1.67-3.07]) and ivermectin (40, 99.9 [70.83-137.18]), with the highest disproportionalities for ivermectin, clarithromycin and niraparib (11, 10.54 [5.24-18.94]).

A total of 10 drug classes were associated with the bradyphrenia cluster (2 unexpected, 2 integrated, 6 expected), the most common drugs being natalizumab (105, 4.65 [3.77-5.67]), levothyroxine (85, 2.97 [2.36-3.7]) and interferon beta-1a (65, 2.6 [2-3.34]), the strongest signals being with lorcaserin (17, 40.85 [23.67-65.71]), finasteride (33, 11.9 [8.16-16.79]) and natalizumab.

Finally, 44 drug classes were associated with the incoherence cluster (34 expected, 4 integrated, 6 unexpected), the more numerous substances being levothyroxine (237, 1.9 [1.67-2.17]), interferon beta-1a (213, 1.98 [1.72-2.27]) and montelukast (200, 5.58 [4.82-6.43]), and the highest disproportionalities being those with anti-infectives –mefloquine (33, 36.26 [24.8-51.28]), zanamivir (14, 11.95 [6.51-20.13]) and oseltamivir (62, 6.47 [4.95-8.31]).

## Discussion

### Overview

Given the increased focus on communication-related markers of affective and psychotic disorders, there is an increased need for a careful overview of how medications could act as confounders. The current study rigorously combined evidence from drug package inserts with post-marketing disproportionality analyses and relied on causal inference techniques to account for potential biases, in order to provide a first attempt at such an overview.

In the following subsections, we discuss how to interpret and use these findings and methods in the broader context of digital phenotyping trying to identify markers of neuropsychiatric conditions: discussing expected and unexpected potential adverse reactions as they relate to the specific context of communication-related markers of psychotic and affective disorders; presenting the limitations and possibilities of our approach; and discussing possible realistic uses of the list in future research.

### Known and unexpected adverse reactions

The final list of potential confounding drugs for communication-related markers encompasses both expected (i.e., described in the package insert) and unexpected associations. Some of the expected associations are already discussed in the literature on communication-related markers. For example, the effects of antipsychotics and antidepressants have been directly investigated when evaluating communication-related markers (Cohen et al., 2017; Cummins et al., 2015; de Boer et al., 2020; Püschel et al., 1998; Sinha et al., 2015). However, even these expected associations are not routinely considered in the actual analysis of communication-related markers of psychotic disorders (Parola et al., 2020), and when they are, the results are inconclusive (Parola, Lin, et al., 2022; Parola, Simonsen, et al., 2022).

In other cases, we found unexpected associations with drugs from already known classes (integrated findings, that is, drugs from the same class were already known to associate with the adverse reaction). For instance, clonazepam (an antiepileptic, also used to treat anxiety) being associated with the aphasia cluster and for antineoplastic agents (mainly VEGFR-inhibitors) with the dysphonia cluster. Some drugs’ package inserts, in fact, did not list all the possible preferred terms that could be used to describe their side effects. This led to the classification of certain expected drug reactions as unexpected (e.g., the package inserts for haloperidol only mentioned motor control disorders, but not speech motor control disorders), even if the scientific literature or clinical practitioners may already be aware of them.

Other associations are more unexpected. Medications used to treat cancer, such as plant alkaloids, cytotoxic antibiotics, protein kinase inhibitors, and monoclonal antibodies, emerge as potential causes of aphasia, which are not reported in the SIDER database. Crucially, since there is at least some evidence of increased cancer risk in schizophrenia (Nordentoft et al., 2021), we could expect a more common use of these drugs in patients with schizophrenia than in controls. Therefore, the adverse reaction could influence how well a predictive model could detect psychotic disorders from speech or language patterns, at least in complex machine learning models. Nevertheless, these drugs have never been mentioned - to our knowledge - in previous studies of communication-related markers as possible confounders.

### Drug-induced communicative impairment mechanisms

We contextualized the drugs identified as possible confounders for communication-related markers according to their plausible mechanism of action (see **Figure 3**). Indeed, biological plausibility is one element of credibility for hypotheses emerging from disproportionality analyses. Furthermore, understanding the mechanism underlying drug-induced communication atypicalities may allow to identify other plausible involved drugs not detected in our study (e.g., because of unaccounted for biases, or because still not on the market). Finally, the knowledge of exactly how communication-related markers are affected by each drug may be included in machine learning algorithm to provide more reliable predictions.

**Figure 3.**
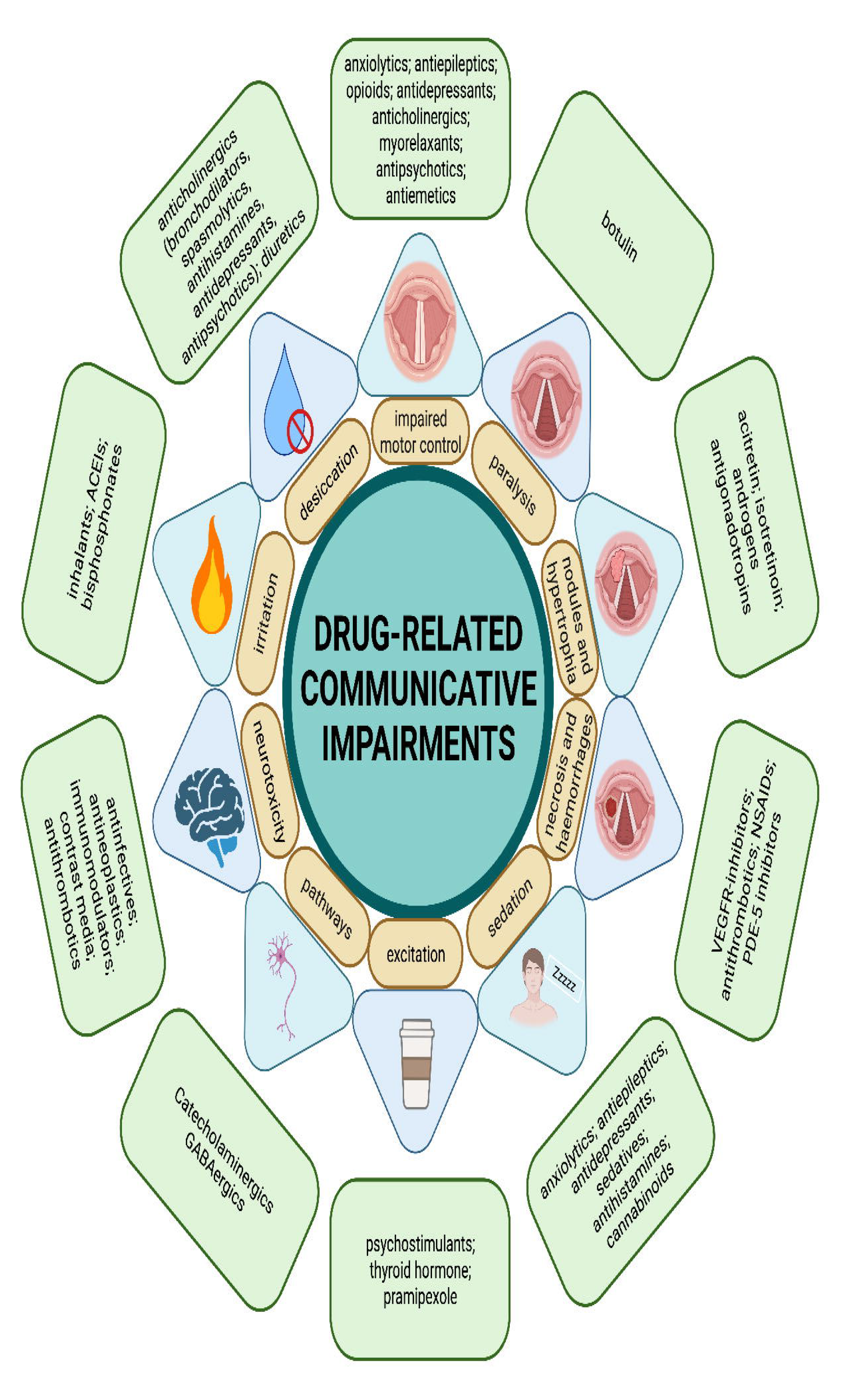
Summary of drug-related communicative atypicalities’ plausible mechanisms. Created with BioRender.com.

The role of drugs in inducing phonatory impairment, often reported as hoarseness, is already consolidated for multiple drugs (see **Table S3**). The primary responsible - in terms of numbers - is plausibly **anticholinergic toxicity** because of xerostomia and larynx desiccation (antimuscarinic inhaled bronchodilators, spasmolytics, drugs for overactive bladder, muscle relaxants, antidepressants, antipsychotics, antihistamines) (Haft et al., 2015). Notably, we observed a signal for second and third generation antihistamines which, trespassing less the blood brain barrier, preserve from central anticholinergic toxicity (mainly sedation) but may nonetheless exert their peripheric effect on salivary glands. The drying effect of diuretics, secondary to hypovolemia, is controversial (Schwartz et al., 2009). Instead, drug-related **laryngeal irritation** is an established common condition, whether because of inhalant drugs (corticosteroids – especially dry powders (Galván & Guarderas, 2012) – beta-agonists and mast-cell stabilizers), drugs inducing cough such as angiotensin converting enzyme inhibitors (Bangalore et al., 2010), or improperly taken bisphosphonates (Hanna et al., 2012). In fact, for inhalants and other respiratory drugs (xanthines, leukotriene receptor antagonists, respiratory monoclonal antibodies), it is often difficult to differentiate between the role of the drug and the underlying disease.

Drug-induced **organic lesions of vocal cords** may also be responsible for dysphonia, as in the case of hemorrhages induced by anti-thrombotics, anti-inflammatories, and 5-phosphodiesterase inhibitors (Stachler et al., 2018), reversible nodules due to excessive granulation response induced by acitretin and isotretinoin (Busso & Serrano, 2005; Kim et al., 2006), or necrosis due to the antiangiogenetic activity of VEGFR-inhibitors (Kudo et al., 2018; Melo et al., 2019; Motzer et al., 2013; Saavedra et al., 2014; Sulibhavi et al., 2020; Wen et al., 2018). Sex hormones may also be involved (Zamponi et al., 2021), as for androgens and antigonadotropins inducing vocal cords thickening and voice deepening through androgen receptors on the larynx (Chadwick et al., 2021). Furthermore, antineoplastics and immunomodulating drugs are also known to be associated with dysphonia, plausibly due both to the cytotoxic (Berretta et al., 2004) and immunomodulating role of the drug (Benfaremo et al., 2018; Bruno et al., 2021), to the disease (Gavrila et al., 2015), and to concomitant radiotherapy (Villari & Courey, 2015).

Finally, an impairment in phonation may be due to extrapyramidal **dystonia** (mainly antipsychotics, but also dopamine antagonist antiemetics such as metoclopramide, that was subjected to an FDA black box warning for dyskinesia, with involuntary movements of the tongue) or to botulinum-related **flaccid paralysis** (a black box warning for systemic toxicity was added to the package insert on 2009).

Other drug classes expected based on SIDER are insulins, 5HT3 antagonist antiemetics, antimycotics, antivirals, dopamine agonists, cholinergic drugs, cough preparations, antiepileptics, analgesics and anesthetics, anxiolytics and sedatives, and cardiovascular drugs. These drug are themselves not totally free of confounding, such as confounding by indication (DAG C: diabetes (Hamdan et al., 2013), cough, and vomit) and reverse causality (DAG B: proton pump inhibitors – for dysphonia supposedly due to laryngo-esophageal reflux (Lechien et al., 2017; Ruiz et al., 2014) – and antibiotics – for dysphonia supposedly due to respiratory infections (Stachler et al., 2018)).

The role of drugs in inducing speech motor control impairment is already consolidated for dopamine antagonists-related acute dystonia and tardive dyskinesia (antipsychotics), agents inducing sedation and reduced speech motor control (anxiolytics, antiepileptics, opioids, antidepressants, anticholinergic drugs, muscle relaxants), neurotoxic drugs (anti-infective, antineoplastic and immunomodulator agents), dopamine agonists (Craig-McQuaide et al., 2014), and drugs interacting with catecholaminergic and GABAergic pathways (Ekhart et al., 2021). We also observed an association with antineoplastics and immunomodulators – plausibly due to their neurotoxicity – and with cardiovascular agents and hormones. Interestingly, the signal for antithrombotic medications persisted when excluding ischemic and unspecified stroke cases. Even if we cannot exclude reverse causality and indication bias, this signal may point to the possibility of drug-induced cerebral hemorrhages.

Multiple cases of iatrogenic aphasia have been reported in the last decade (Rizwan et al., 2021), often concerning reversible conditions induced by immunomodulators, chemotherapy and fluoroquinolones-related neurotoxicity (Belin et al., 2020; Bennett et al., 2019; Carl et al., 2015; Higa et al., 1995; Patel et al., 2015). A similar toxicity may also manifest because of the increased permeability of the blood brain barrier due to contrast media, potentially allowing endogenous and exogenous neurotoxins to reach the central nervous system. Dopamine antagonism (Chien et al., 2017), shared by antipsychotics and the propulsive domperidone, may also be responsible for aphasia, as well as antithrombotic-related hemorrhages. Bradyphrenia and tachyphrenia may also be the manifestation of neurotoxicity, and of sedation (e.g., antiepileptics, pramipexole, antipsychotics, lithium, benzodiazepines, antidepressants, antihistamines, cannabinoids) and excitation (levothyroxine, psychostimulants), respectively.

### Limitations and future directions

#### Formalized query

In the attempt to retrieve cases of interest in the FAERS, we found an often-ambiguous lexicon covering communicative impairments. The current study explicitly formalized a MedDRA query to retrieve communicative impairments relevant to affective and psychotic disorders more systematically. This formalized query is a necessary step to focus the attention and to create a common framework for disproportionality analyses on these impairments.

The current query presents some limitations. First, one might more closely investigate how physicians describe and report these impairments. For example, common terms used by physicians to report dysphonia are acute laryngitis, nonspecific dysphonia, benign vocal fold lesions, and chronic laryngitis (Stachler et al., 2018), and for retrieving antipsychotic-related dysarthria cases one may search also for extrapyramidal syndrome and laryngospasm. More work is needed to cover these labels and validate the results of searches that integrate them. Second, perhaps more crucially, we observed a high proportion of communication-related FAERS cases submitted by the general public. This suggests that communicative adverse events might be at the same time underplayed by medical practitioners, and of crucial importance to patients, caregivers, and families. In fact, we observed that patients with communicative impairments tend to specify the resulting disability more frequently in their reports than patients with other adverse events but the same underlying condition. Third, during the definition of the query, we found several inconsistencies between the clusters of relevant MedDRA terms and terminological practices in the speech and language pathology community, which could create unnecessary confusion.

One could also question whether FAERS’ and SIDER information is sufficiently sensitive to the kind of properties analyzed in the search for communication-related markers. For example, minor acoustic atypicalities such as increased jitter – low-level irregularities in voice pitch, a commonly used acoustic measure in predictive, machine learning algorithms for affective and psychotic disorders (Cummins et al., 2015; Parola et al., 2020) as well as for Parkinson’s disease (Tsanas & Arora, 2021) – might not be perceived, or at least not perceived as enough of an issue, by patients and clinicians to be reported and precisely labeled. This suggests that a closer collaboration of patients and practitioners – crucially including speech and language pathology and communicative markers experts – in developing a common and easy to use terminology and clear definitions for communicative impairments would provide a substantial improvement for the MedDRA lexicon.

Nevertheless, the construction of an initial query enables initial explorations of medication-based confounders and facilitates proposals, thus representing an important step in the development of a useful Standardized MedDRA Query (SMQ).

#### Causal inference

Although still uncommon in disproportionality analyses, formalized causal inference, and the use of DAGs, in particular, are a promising endeavor (Cunningham, 2021; Pearl, 2009). These tools provide a standardized framework for the formalization, visualization, and communication of confounding. These tools also provide structured and more reproducible procedures to account for at least some of the biases when designing analyses (Pearl, 2009).

We have built four relatively simple DAGs of the mechanisms underlying observed drug event observations. Thus, we have tried to identify the most problematic biases for our questions and accordingly adjusted our analyses and interpretation. However, it is important to note that many biases could not be fixed and that the characteristics of the reporting (often incomplete and unverified) complicate attempts at causal inference. For example, proton pump inhibitors are used to treat or prevent gastroesophageal reflux, a condition that may also affect the larynx and vocal cords and result in laryngo-esophageal reflux disease and dysphonia (Lechien et al., 2017). Therefore the causal direction (PPI to dysphonia, or reflux to both PPI and dysphonia) cannot be easily identified. Further, our broad focus did not permit us to delve into the richness of spontaneous reports (e.g., information on concomitants, therapy regimen, co-occurring events) and to map more complex scenarios (e.g., variables affecting at the same time the use of the drug, the incidence of the adverse event, and the reporting of it). For example, botulinum toxin has been referred to as a potential cause and treatment for spasmodic dysphonia but may also temporarily cause dysphonia through muscle weakening. In addition, biases, such as notoriety bias, and masking bias, adjustment for the Weber effect (Raschi et al., 2018), are beyond the scope of this study but should be considered when investigating specific drugs more closely. Further, because of the many biases of spontaneous reporting, the comparison of the safety profile of different drugs on the basis of disproportionality alone tends to be unreliable and is in general not recommended (Mouffak et al., 2021).

#### Integrating additional sources

The main objective of spontaneous reporting systems is to collect useful data to identify unexpected associations between a drug and an adverse event in a timely and cost-effective manner. This identification enables early intervention and therefore limits the costs of drug-related harm. To effectively target currently not known safety issues, it is extremely important to integrate already acquired knowledge, which may come from the literature, or from regulatory sources - primarily package inserts (FDA) and Summaries of product characteristics (EMA).

Databases that store this information in an easily accessible way are a promising tool for large-scale analyses because reading and coding each individual package insert would be extremely time consuming. The SIDER uses a natural language processing algorithm to extract the information from regulatory sources and has not been updated since 2016 (Kuhn et al., 2016), therefore, it plausibly contains errors and outdated information. Furthermore, it may contain terms linked but not coincident with the investigated events (as in the case of haloperidol), and therefore our automated process may lack some expected reactions.

A worldwide database in which data for each marketed drug is compiled and regularly updated by the marketing authorization holder and stored in an accessible way would enrich both regulatory activities and disproportionality analyses. In the meantime, the use of the SIDER or similar databases may help in large-scale analyses to reduce the risk of classifying already known reactions as unexpected signals.

We cannot be sure whether some of the unexpected associations have already appeared as notes in clinical trials (but not reported in the package insert) or in subsequent scientific literature. Future work should attempt to integrate these additional sources of information. However, independent of the novelty, our list aggregates large amounts of otherwise dispersed information in an easier to consult format.

Future work could integrate additional sources of information, both as related to known associations (e.g., scientific literature) and additional clinical data (e.g., health records), to provide a more comprehensive overview. Further, different sources could be weighted according to the degree of evidence available (e.g., via Bayesian analysis).

#### Large-scale analyses

Traditional disproportionality analyses focus on at most a handful of drugs and/or adverse events (Aiello et al., 2021; Raschi et al., 2020). Thus, they can provide a fine-grained analysis of potential confounders, including a nuanced analysis of how sociodemographic variables might affect drug prescription and adverse reactions (Hoekstra et al., 2021).

Large-scale analyses require a broader overview, which cannot match the same level of detail and discussion. The strategies we implemented to simultaneously assess large sets of adverse events and drugs may help design future large-scale analyses. These strategies range from correction for multiple testing and automatic integration with regulatory databases, to an attempt to formalize possible underlying causal mechanisms and the use of a priori expected biases to implement robustness analyses. Large-scale analyses, however, provide only an initial perspective and must be complemented with more detailed studies of specific associations and their confounds.

#### How should this list be used?

We advocate for the list of drug confounders (Figure 3 and Supplementary Material 1 – Section D – Table S10)– whether as a cause of communicative atypicality or as a proxy of an underlying susceptibility – to be used in future studies of communication-related behavioral markers by either including the presence of a medication as a covariate, removing participants who take medication, or interpreting results and study limitations as a function of which medications were taken. As observed in multiple reviews, most studies of such markers involve small sample sizes (R. Fusaroli et al., 2017; Parola et al., 2020; Weed & Fusaroli, 2020). Such studies would be at a loss trying to adjust for such a large number of medications and would lack reliable evidence related to all but the most commonly used ones (Rocca & Yarkoni, 2021; Westfall & Yarkoni, 2016). Although a single study may still check the list to identify likely cautions (e.g., much higher use of drug x in the target population than in the controls), the real potential lies in the cumulative aggregation of this information across studies. The key is to promote transparency of reporting and record medications used by participants in individual studies, which would allow future mega-analyses (R. Fusaroli et al., 2022) (aggregating datasets across studies preserving individual-level data) to directly assess the impact of a large variety of relevant medications.

Accounting for confounders is also important in machine learning studies. Current reviews and perspectives on the study of communication-related behavioral markers advocate the collection of larger and more diverse samples and the use of state-of-the-art machine learning techniques, such as deep learning (Parola, Lin, et al., 2022; Parola, Simonsen, et al., 2022; Rybner et al., 2022). In these contexts, the algorithms can detect even the presence of weak confounding if it improves prediction. In other words, many machine learning models are likely to recognize small differences between groups they try to classify. If these differences are due to higher levels of medication being used and not due to the target disorder, the models may not generalize well to other samples of the disorder where the medication use is different, which is common when changing countries and sociodemographic settings. Accordingly, a deeper understanding of the confounders and mechanisms at work is a key component also for more data-driven machine learning approaches, for instance, to guide bias assessment or even to identify more rigorous pipelines (e.g., presenting medication-balanced validation sets).

Finally, this list may also help identify more general hypothesized mechanisms underlying adverse events beyond a specific drug. Pharmacosurveillance can thus act not only as a guide for precautionary regulatory action, but also as a hypothesis generation tool for scientific research, which could lead to follow-up studies involving, e.g., electronic health records (to assess adverse events before and after drug administration), experimental setups, and clinical studies. For instance, a more thorough investigation of the association between domperidone and aphasia would be of particular interest, given the biological plausibility (i.e., its activity as a dopamine antagonist) and the existence of conditions that increase the blood-brain barrier permeability. This might lead to more generalizable predictions regarding confounding drugs and an increased understanding of the communicative features of the disorders over time.

### Applications of the methods to other neuropsychiatric conditions

In the current study, we have focused on affective and psychotic disorders since previous research explicitly called for better investigation of medication-related confounders in identifying communication markers for these populations (Cummins et al., 2015; Low et al., 2020; Parola et al., 2020). However, with proper consideration, the list could be easily extended when assessing communication-related behavioral markers for other conditions such as neurodevelopmental (e.g., autistic spectrum disorder) and neurological (e.g., Parkinson’s disease) disorders. In particular, for Parkinson’s disease, good practices to account for med-on and med-off levodopa state already exist (e.g., Im et al., 2019; Thies et al., 2021).

## Conclusions

Motivated by the increasing interest in communication-related behavioral markers of affective and psychotic disorders, we set out to investigate the potential role of medications in affecting communication-related markers of these disorders. We extracted the drugs already expected to cause communicative impairments from the SIDER. This paved the way for a pharmacovigilance analysis of a larger set of communication-related adverse events and drugs, controlling for prominent biases.

We identified a list of medications to be accounted for in future studies on communication and biobehavioral markers of affective and psychotic disorders. These studies should take into account: drugs irritating vocal cords and the larynx (inhalants, bisphosphonates, angiotensin-converting enzyme inhibitors); drugs inducing laryngeal desiccation (anticholinergics, diuretics); drugs impairing speech motor control (anxiolytics, antiepileptics, opioids, antidepressants, anticholinergics, myorelaxants, antipsychotics, antiemetics), or temporarily paralyzing vocal cords (botulinum toxin); drugs inducing laryngeal hypertrophy (androgens, antigonadotropins) or the development of nodules on vocal cords (retinoids); drugs potentially inducing necrosis (VEGFR-inhibitors) or hemorrhages in the vocal cords (antithrombotics, nonsteroidal anti-inflammatories, PDE5-inhibitors); sedatives (anxiolytics, antiepileptics, antidepressants, hypnotics, antihistamines, cannabinoids); stimulants (psychostimulants, thyroid hormones, pramipexole); drugs interacting with catecholaminergic and GABAergic neurotransmitters; neurotoxic drugs (antiinfectives, antineoplastics, immunomodulators, contrast media, antithrombotics).

The work showcases methodological innovations to facilitate large-scale disproportionality analyses and identifies current shortcomings, along with discussing potential causal and pathogenetic mechanisms. In particular, the existing lexicon to identify communicative adverse events is ambiguous and not well defined, perhaps due to an underappreciation of the perspectives of patients and lack of integration of experts in speech and language pathology and in communicative impairments. We advocate for future work on this.

Drugs that confound the effect between communication-related behavioral markers and psychiatric disorders are abundant. There should be concern not only for confounding drugs and comorbidities, but also non-medical substances and habits (e.g., smoking, vocal use). Here, we provide a tool for learning about and potentially adjusting for the confounders to improve digital phenotyping research.

## Declarations

## Ethics

Anonymized data were collected from a publicly available database and did not require ethics committee approval.

## Authors’ contributions

MF, AS, and RF conceived the research project. MF, SB, AS, DML, AP, and RF defined the query for case retrieval. MF designed and executed the statistical analysis. MF, ER, EP, and RF reviewed and critiqued the statistical analysis. All the authors contributed to the interpretation of data. Regarding manuscript preparation, MF and RF executed the writing of the first draft and all the authors contributed to the review and critique. All authors approved the final version.

## Data Availability

All data used in the present study are available online at https://fis.fda.gov/extensions/FPD-QDE-FAERS/FPD-QDE-FAERS.html. The data produced in the present study are available upon reasonable request to the authors.

https://osf.io/e374k/

## Acknowledgments

MF, EP and ER were supported by institutional research funds (Ricerca Fondamentale Orientata). AS was supported by a Postdoctoral Fellowship from the Carlsberg Foundation. AP was supported by a Marie Curie Fellowship from the ERC. DML was supported by a RallyPoint Fellowship. MedDRA® trademark is registered by ICH.

## Data availability statement

The pharmacovigilance data are freely accessible on the FDA website: https://fis.fda.gov/extensions/FPD-QDE-FAERS/FPD-QDE-FAERS.html

## Code access

The code used is available upon request.

## BIBLIOGRAPHY

### Supplementary Material

Supplementary Material 1 includes an overview of the studies assessing the effect of medication on speech patterns in schizophrenia (Section A), the MedDRA Query developed for the retrieval of communication atypicalites (Section B), the features of the population investigated (Section C) and the summary of the results of the disproportionality analysis (Section D).

Supplementary Materia 2 stores all the results of the disproportionality analysis, including the effect sizes.

## Footnotes

1 While the adverse event is causing the prescription of the drug, the drug itself could be affecting the symptom (e.g., diminishing it) and therefore a more nuanced causal model than this simplified DAG would have to include bidirectional causal arrows, or a temporal dimension to causation.

