## Supplemental Material 1 for "Identifying medications underlying communication atypicalities in psychotic and affective disorders: A pharmacovigilance study within the FDA Adverse Event Reporting System"

### Supplementary Material 1

- Section A – Overview of the studies assessing the effect of medication on speech patterns in schizophrenia
- Section B – MedDRA Query developed for the retrieval of communicative atypicalities reports
- Section C – Population Investigated
- Section D – Disproportionality and Robustness Analyses

### Supplementary Material 2

- Reporting Odds Ratios

### Section A – Overview of the studies assessing the effect of medication on speech patterns in schizophrenia

The literature to date evaluating the role of medication on speech patterns in schizophrenia is very sparse. A very limited number of studies (n = 5) explicitly took into account the role of antipsychotic medications in influencing speech patterns:

- Puschel et al. (1998) compared acoustic patterns between patients with schizophrenia and control subjects and examined the effect of medication on vocal features. The authors found an association between medication effect, i.e., "neurological side effects of treatment," and the speech parameters "total recording time," "total length of pauses," and "mean pause duration per second"; however, the effects were in an unexpected direction (higher symptoms associated with longer pauses), whereas the other medication effects were not associated with the acoustic parameters. The authors therefore concluded that "Acute medication effects did not explain these findings".

- Cannizaro et al. (2005) studied 13 patients with schizophrenia who participated in a pharmaceutical clinical trial and were treated with a clinically effective dose of risperidone for 14 days. The authors compared acoustic features using two speech tasks of varying complexity (free speech vs. constrained task, i.e., reading) to determine distinct and overlapping features of speech pauses. The authors found that participants with schizophrenia in the free speech condition used greater total pause and a higher percentage of pauses, as well as longer pauses and greater variation in pauses in both the constrained- speech task and the free speech task.
- Cohen et al. (2017) examined the speech patterns of patients with schizophrenia who participated in a six-week randomized controlled trial in which galantamine or oxytocin was administered. The assessment was conducted at baseline and repeated six weeks later. The authors did not find significant change in voice or facial expressions as a result of either galantamine or oxytocin treatments.
- de Boer et al. (2020) compared patients with schizophrenia who received antipsychotic medications divided into two categories based on their mechanism of action, i.e., high and low dopamine occupancy of the D2 receptor (D2R), respectively. Overall, they found more severe negative speech disorders (i.e., slower articulation rate, increased pauses, and shorter utterances) in patients taking antipsychotics with high D2R occupancy, whereas patients with low D2R occupancy had less pronounced disorders, suggesting that speech disorders may be exacerbated by antipsychotics with high D2R occupancy.

- Parola et al. (2022) sought to assess the generalizability of de Boer et al.'s (2021) findings in a cross-linguistic sample of 231 patients with schizophrenia and 238 matched controls in four languages (Danish, German, Mandarin, and Japanese). The authors found that medication type (high or low D2 receptor occupancy) was related to voice patterns, but inconsistently across languages. The most prevalent patterns were that patients taking medication with high D2R occupation had lower pitch variability, higher number of pauses, longer turn duration, and higher speech rate compared to patients taking medication with low D2R occupation. Higher medication dosage (CPZ equivalents) was associated with lower median pitch, lower speech rate, longer pause and utterance duration, and lower total number of words.

Overall, the few studies available, the small sample sizes, the limited information on the characteristics of drug treatment, and the large differences between studies do not allow conclusions to be drawn about the effects of medication on speech patterns in schizophrenia. The only potentially reliable effect indicated by the studies to date which need to be further explored in future studies is the difference found in the studies by de Boer et al. (2020) and Parola et al. (2022) between high and low D2R medications.

### Section B – MedDRA Query developed for the retrieval of communicative atypicalities reports

**Table S1 – MedDRA Query** for the retrieval of communicative atypicalities reports. We identified multiple sub-queries including semantically overlapping MedDRA Preferred Terms. The clusters were obtained on the basis of semantic overlapping, and therefore on the possibility that the reporters may have used interchangeably different terms, rather than with a reference to existing speech and language pathologies.

| **Cluster** | **MedDRA Preferred Terms** | | |
|---|---|---|---|
| **Cluster 1 (dysphonia-related)** | Dysphonia<br>Dysphonia psychogenic<br>Muscle tension dysphonia | Spasmodic dysphonia<br>Aphonia<br>Aphonia psychogenic | Phonastenia<br>Stridor |
| **Cluster 2 (speech motor control-related)** | Dysarthria<br>Dyslalia | Dysphemia | |
| **Cluster 3 (prosody-related)** | Aprosody<br>Dysprosody | | |
| **Cluster 4 (aphasia-related)** | Aphasia<br>Primary progressive aphasia | | |
| **Cluster 5 (tachyphrenia-related)** | Logorrhea<br>Pressure of speech | Flight of ideas<br>Tachyphrenia | |
| **Cluster 6 (bradyphrenia-related)** | Poverty of speech<br>Bradyphrenia | Lack of spontaneous speech<br>Poverty of thought content | Taciturnity<br>Thought blocking |
| **Cluster 7 (abnormal reasoning-related)** | Ideas of reference<br>Illogical thinking | Impaired reasoning<br>Magical thinking | Paralogism |
| **Cluster 8 (stereotypy-related)** | Coprolalia<br>Echolalia | Perseveration<br>Repetitive speech | Verbigeration |
| **Cluster 9 (incoherence-related)** | Disorganized speech<br>Incoherent<br>Clang associations | Derailment<br>Loose associations<br>Tangentiality | Thinking abnormal |
| **Other terms excluded from clusters** | Pedantic speech<br>Intellectualization<br>Morbid thoughts<br>Pathological doubt<br>Intrusive thoughts<br>Circumstantiality | Speech disorder developmental<br>Mutism<br>Speech disorder<br>Cognitive linguistic disorder | Social communication disorder<br>Language disorder<br>Speech sound disorder<br>Slow speech<br>Confabulation |

#### Section C – Population investigated

Using the query shown in the Table S1, we extracted from the deduplicated FAERS the cases of interest (any communication impairment), separated by underlying condition (psychotic, affective, and non-neuropsychiatric reports –i.e., reports without neurologic drugs; **Figure S1**).

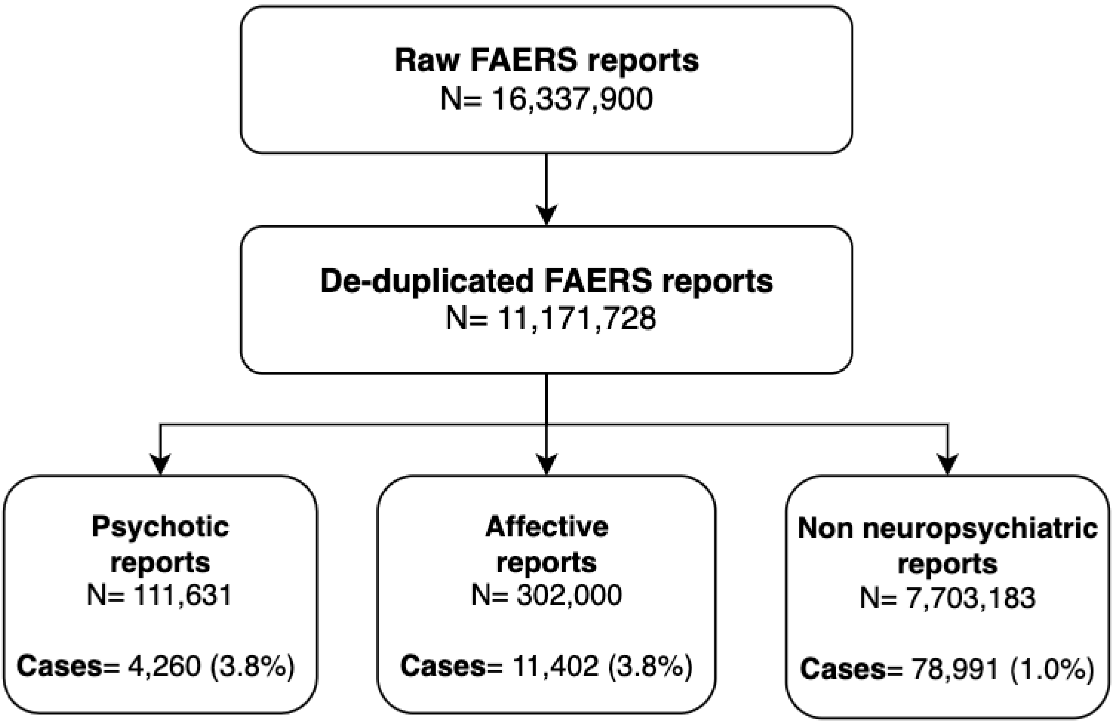

**Figure S1 – Case retrieval flowchart.** Cases (i.e., any communication-impairment) and non-cases in the three populations as retrieved from the FAERS. Note that some reports can be included in more than one population. 12,319 reports (599 cases, 11,720 non-cases) were included in both the psychotic and the affective populations.

To identify potential risk factors or biases, we compared demographic and reporting information between cases and non-cases in each population (Table S2).

Most of the reports (>80%) were submitted from North America and Europe. Consumers submitted more communicative-impairment reports within psychotic and non-neuropsychiatric reports. The numbers of co-reported comorbidities and concomitant drugs were similar between cases and non-cases. However, the presence of communicative impairments was associated, across the three populations, with a higher number of co-occurrent adverse events (i.e., more symptoms are recorded in each report), disability, and hospitalization rate.

**Table S2 – Demographic characterization of the reports**. For each population, we compared frequency of reports (absolute number and valid percentage –i.e., not including missing values), sex and age, continent of reporting, role of the reporter, outcome, and number of co-occurrent symptoms.

| | Psychotic disorders | | | | Affective disorders | | | | Non-neuropsychiatric | | | |
|---|---|---|---|---|---|---|---|---|---|---|---|---|
| | Cases | | Non-cases | | Cases | | Non-cases | | Cases | | Non-cases | |
| | N | % | N | % | N | % | N | % | N | % | N | % |
| **Reports** | 4,260 | 3.8 | 107,371 | 96.2 | 11,402 | 3.8 | 290,598 | 96.2 | 78,991 | 1.0 | 7,624,192 | 99.0 |
| **Sex** | | | | | | | | | | | | |
| Female | 1,659 | 41.0 | 41,698 | 41.0 | 7,421 | 68.0 | 185,864 | 67.0 | 48,706 | 64.7 | 4,267,049 | 61.6 |
| Male | 2,415 | 59.0 | 59,321 | 59.0 | 3,509 | 32.0 | 89,666 | 33.0 | 26,567 | 35.3 | 2,658,172 | 38.4 |
| Missing | 186 | - | 6,352 | - | 472 | - | 15,068 | - | 3,718 | - | 698,971 | - |
| **Age category** | | | | | | | | | | | | |
| <18 | 95 | 2.9 | 2,660 | 3.4 | 383 | 4.2 | 10,920 | 5.0 | 2,618 | 4.7 | 295,418 | 5.9 |
| 18-29 | 634 | 19.0 | 13,727 | 17.6 | 906 | 10.0 | 23,537 | 11.0 | 2,750 | 5.0 | 383,547 | 7.7 |
| 30-49 | 1,225 | 37.0 | 30,010 | 38.4 | 3,008 | 33.0 | 71,307 | 32.0 | 11,298 | 20.5 | 1,048,614 | 21.0 |
| 50-64 | 751 | 23.0 | 18,590 | 23.8 | 2,818 | 31.0 | 67,206 | 31.0 | 16,808 | 30.4 | 1,515,597 | 30.3 |
| 65-74 | 316 | 9.5 | 7,115 | 9.1 | 1,139 | 13.0 | 27,117 | 12.0 | 11,501 | 20.8 | 875,729 | 19.5 |
| 75-84 | 220 | 6.6 | 4,690 | 6.0 | 605 | 6.7 | 14,674 | 6.7 | 7,903 | 14.3 | 601,304 | 12.0 |
| >84 | 70 | 2.1 | 1,365 | 1.7 | 175 | 1.9 | 5,002 | 2.3 | 2,335 | 4.2 | 183,242 | 3.6 |
| Missing | 949 | - | 29,214 | - | 2,368 | - | 70,835 | - | 23,778 | - | 2,620,741 | - |
| **Reporter** | | | | | | | | | | | | |
| Consumer | 1,474 | 36.0 | 25,546 | 25.0 | 5,245 | 51.0 | 132,509 | 50.0 | 42,718 | 57.5 | 3,512,207 | 48.4 |
| Physician | 1,239 | 31.0 | 39,039 | 38.0 | 2,290 | 22.0 | 66,128 | 25.0 | 14,820 | 20.0 | 1,716,345 | 23.7 |
| Pharmacist | 265 | 6.5 | 9,501 | 9.3 | 452 | 4.4 | 11,834 | 4.5 | 10,052 | 13.5 | 1,102,747 | 15.2 |
| Lawyer, healthcare practitioner or other | 1,078 | 26.5 | 28,334 | 27.7 | 2,199 | 22.6 | 55,101 | 20.5 | 6,683 | 9.0 | 918,824 | 12.7 |
| Missing | 204 | - | 4,951 | - | 1,216 | - | 25,026 | - | 4,718 | - | 374,069 | - |
| **Continent** | | | | | | | | | | | | |
| North america | 1,965 | 48.0 | 49,878 | 49.0 | 7,243 | 68.0 | 192,321 | 70.0 | 57,201 | 48.0 | 48,878 | 48.5 |
| Europe | 1,543 | 38.0 | 37,129 | 36.0 | 2,479 | 23.0 | 60,169 | 22.0 | 11,192 | 37.7 | 37,129 | 36.1 |
| Asia | 304 | 7.4 | 9,112 | 8.9 | 437 | 4.1 | 12,207 | 4.4 | 4,183 | 7.4 | 9,112 | 8.9 |
| Other | 282 | 6.6 | 6,631 | 6.1 | 491 | 4.9 | 9,713 | 3.6 | 3,828 | 6.9 | 6,631 | 6.5 |
| Missing | 166 | - | 4,621 | - | 752 | - | 16,188 | - | 2,587 | - | 4,621 | - |
| **Outcome** | | | | | | | | | | | | |
| Death or life-threat | 492 | 11.5 | 18,758 | 17.5 | 1,146 | 10.1 | 27,729 | 9.5 | 6,742 | 8.5 | 280,865 | 3.7 |
| Disability or congenital anomaly | 180 | 4.2 | 2,201 | 2.0 | 764 | 6.7 | 12,636 | 4.3 | 3,397 | 4.3 | 129,814 | 1.7 |
| Hospitalization or intervention required | 1,722 | 40.4 | 34,241 | 31.9 | 3,605 | 31.6 | 70,739 | 24.3 | 19,966 | 25.3 | 1,466,799 | 19.2 |
| Other serious | 1,109 | 26.0 | 30,305 | 28.2 | 3,314 | 29.1 | 81,271 | 28.0 | 20,186 | 25.6 | 1,929,892 | 25.3 |
| Non specified outcome | 757 | 17.8 | 21,886 | 20.4 | 2,573 | 22.6 | 98,223 | 33.8 | 28,700 | 36.3 | 3,256,211 | 42.7 |
| **Co-occurrent events** | | | | | | | | | | | | |
| 50% (25%-75%) | 7 (5-11) | | 3 (1-5) | | 8 (5-14) | | 3 (2-5) | | 5 (3-9) | | 2 (1-4) | |
| **Concomitant drugs** | | | | | | | | | | | | |
| 50% (25%-75%) | 4 (2-8) | | 3 (1-6) | | 5 (2-9) | | 4 (2-9) | | 2 (1-4) | | 2 (1-3) | |
| **Comorbidities** | | | | | | | | | | | | |
| 50% (25%-75%) | 3 (1-5) | | 2 (1-4) | | 3 (1-6) | | 3 (1-6) | | 1 (1-2) | | 1 (1-2) | |

There were differences in the adverse events co-reported with cases, relative to non-cases, in the three populations (Figure S2-S5), with anxiety symptoms, consciousness disturbances, dyskinesias and confusion being more frequent in psychotic and affective cases, while multiple respiratory conditions, pain, headache, nausea, and vomit were more common in non-neuropsychiatric reports. Indications and concomitant drugs overlapped between cases and non-cases in affective and psychotic reports, but non-neuropsychiatric ones had a higher proportion of cases with, as indication, multiple sclerosis acute and progressive (12% vs 0%), bronchospasm and obstruction (8% vs 1%) and rheumatoid arthropathies (5% vs 0%), and, as drugs, immunosuppressant agents and selective beta-2 adrenoceptors.

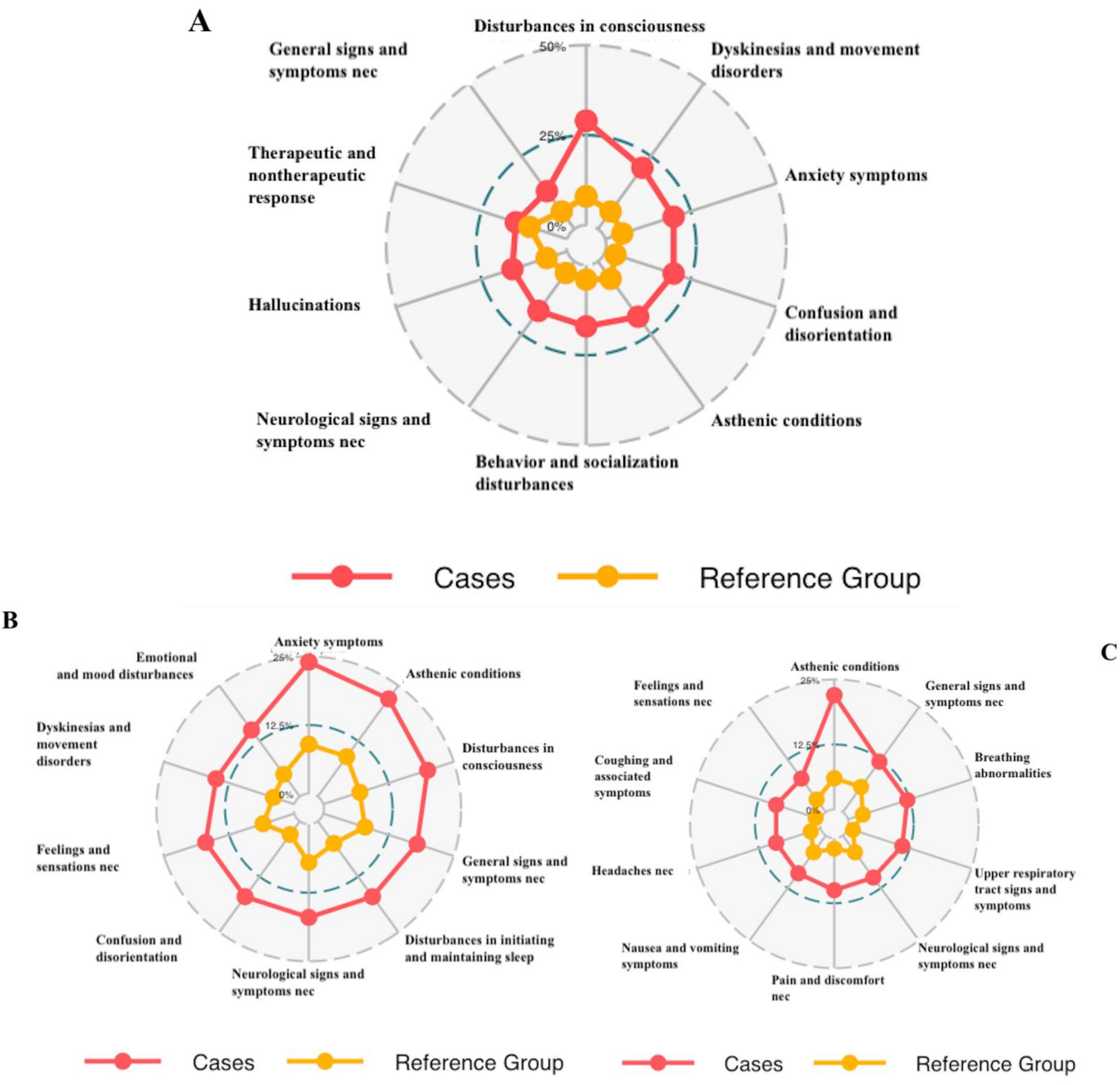

**Figure S2 – Radar-plot of adverse events co-reported with communication ones**.
For each population (A: psychotic, B: affective and C: non-neuropsychiatric reports), we identified the 10 most commonly co-reported adverse events (5 for cases and 5 for non-cases) and displayed in a clockwise order starting from the top. In red, we report the relative incidence of each co-reported adverse event in reports mentioning communication-related adverse events. In yellow we report the non-cases (reference group). To facilitate interpretation, we represent two reference levels (circles): 12.5% and 25% of co-occurrences. Note that, for clarity, panel A has a different scale (25% and 50%) than the other panels.

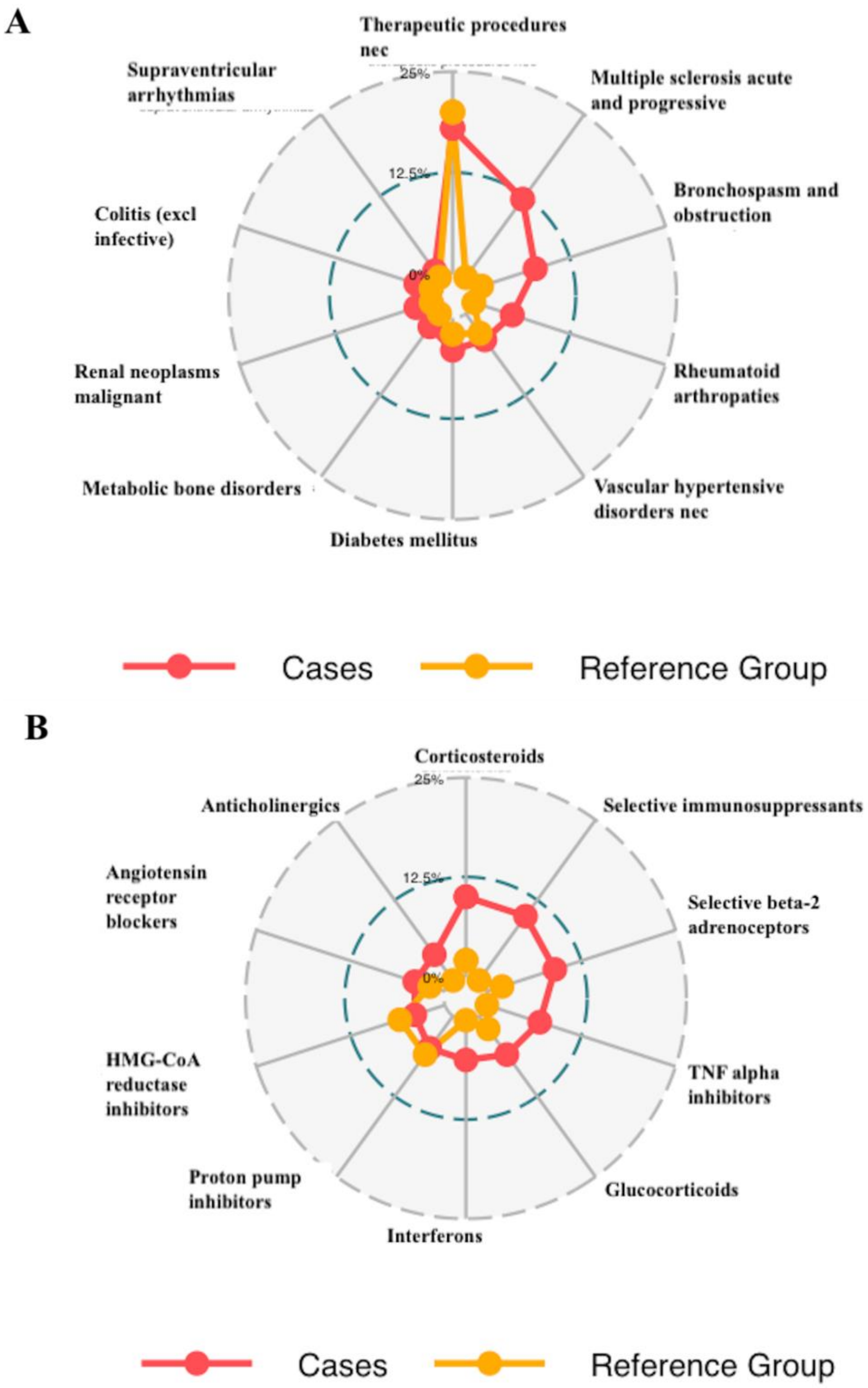

**Figure S3 – Radar-plot of comorbidities and concomitants in non-neuropsychiatric reports**
For non-neuropsychiatric reports, we displayed the 10 most commonly co-reported indications (A) and drugs (B): 5 for cases and 5 for non-cases. We displayed them in a clockwise order starting from the top. In red, we report the relative incidence in reports mentioning communication-related adverse events. In yellow we report the non-cases (reference group). To facilitate interpretation, we represent two reference levels (circles): 12.5% and 25% of co-occurrences.

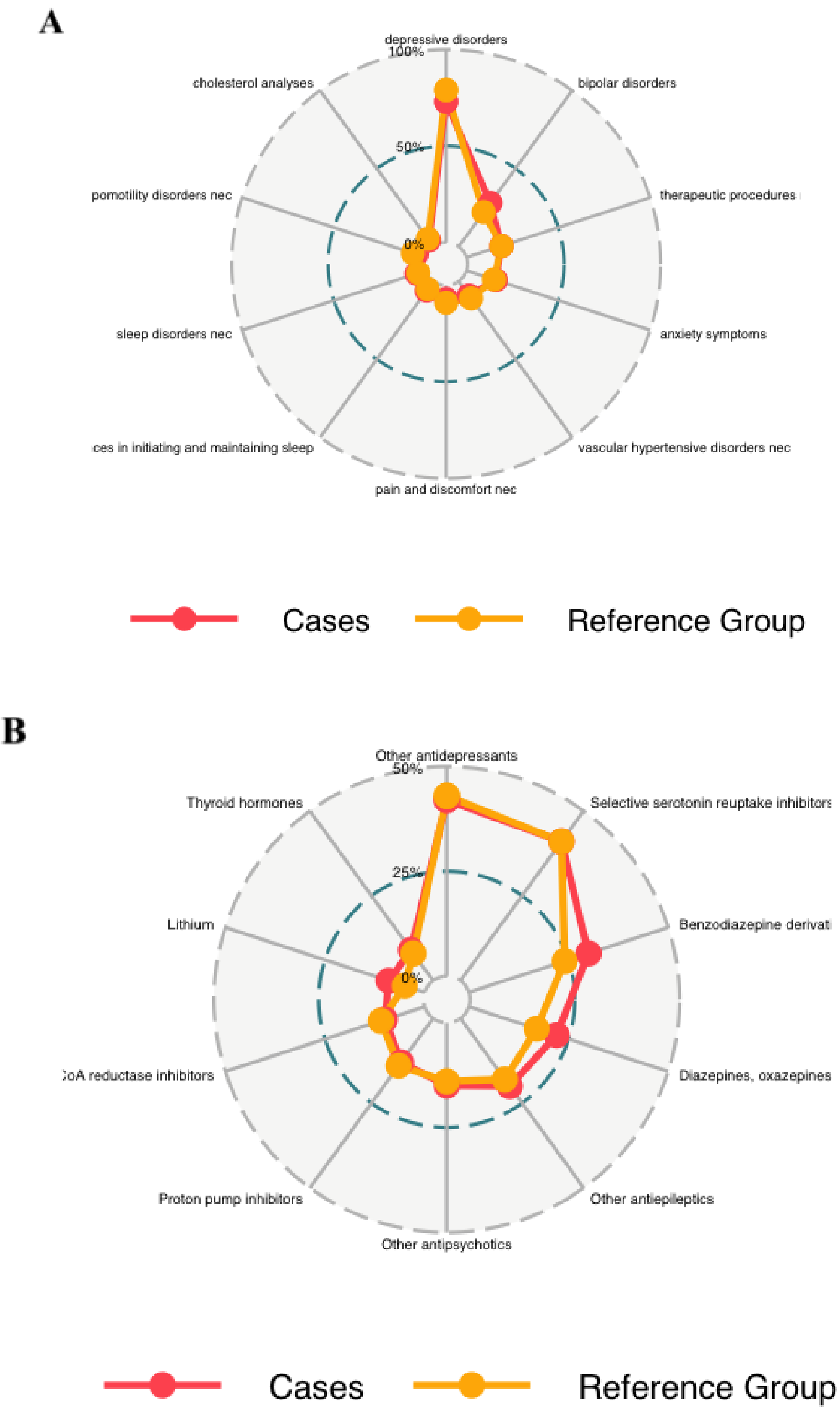

**Figure S4 – Radar-plot of comorbidities and concomitants in affective reports**

For affective reports, we displayed the 10 most commonly co-reported indications (A) and drugs (B): 5 for cases and 5 for non-cases. We displayed them in a clockwise order starting from the top. In red we report the relative incidence in reports mentioning communication-related adverse events. In yellow we report the non-cases (reference group). To facilitate interpretation, we represent two reference levels (circles): 50% and 100% of co-occurrences for A, 25% and 50% of co-occurrences for B.

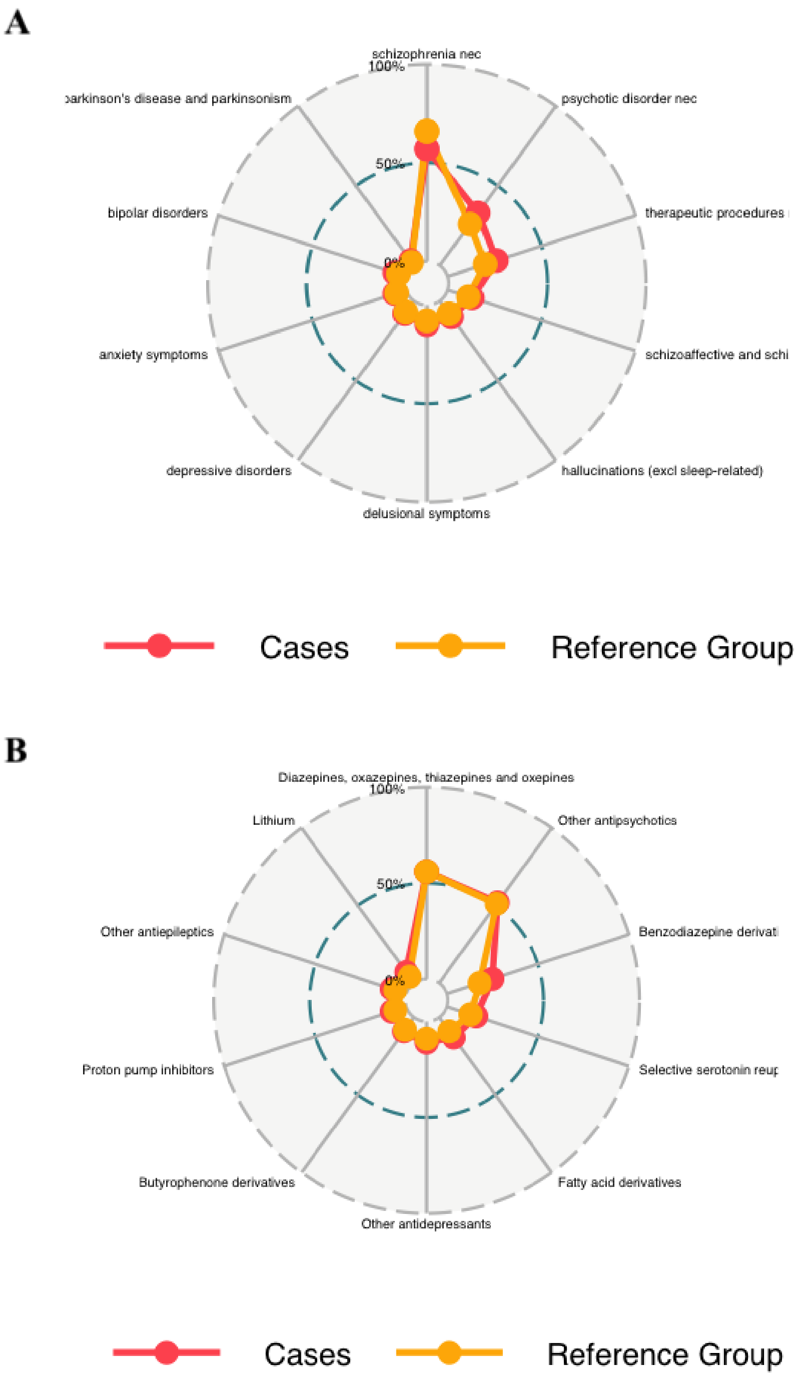

**Figure S5 – Radar-plot of comorbidities and concomitants in psychotic reports**
For psychotic reports, we displayed the 10 most commonly co-reported indications (A) and drugs (B): 5 for cases and 5 for non-cases. We displayed them in a clockwise order starting from the top. In red we report the relative incidence in reports mentioning communication-related adverse events. In yellow we report the non-cases (reference group). To facilitate interpretation, we represent two reference levels (circles): 50% and 100% of co-occurrences.

#### Section D – Disproportionality and Robustness Analyses

We performed a disproportionality analysis evaluating associations between drugs (from the ATC 2022 classification, excluding mineral supplements and drugs included in the 'Various' class) and communication-related adverse events (sub-clusters of overlapping terms as identified in Table S1). The estimated reporting odds ratios (ROR) were shown in Supplementary Materials 2.

For each sub-query, we report a potential association when the adverse event is included in the package insert of the drug (SIDER) or is more likely to be reported together with the drug of interest than with any other drug but the one analyzed (FAERS).

Unexpected drugs, i.e., drugs associated with the sub-queries investigated but not included in the SIDER, were stratified according to expected biases (Figure 2 – Step 4) through clinical reasoning and according to the causal inference framework discussed in the introduction (paragraphs 1.3 and 1.4). In particular, we scanned the unexpected association for potential reverse causality (DAG B), potential confounding by indication (DAG C), and by concomitant (DAG D), and therefore identified by exclusion the uncontroversial ones (plausible adverse reactions, DAG A, for which no specific confounder was expected).

Finally, we performed robustness analyses to partly account for these biases:

- we excluded reports with the specific communicative impairment among indications, or restricted the investigation to a specific indication, to account for reverse causality bias (DAG B; Robustness Analysis 1).
- we excluded reports with pathologies that may be responsible for indication bias (DAG C; Robustness Analysis 2), at least for drugs that are approved for multiple indications (otherwise, the bias was considered non-fixable).
- we excluded reports with the drug responsible for the ambiguity to account for the concomitant bias (DAG D; Robustness Analysis 3).

Here we present the results of the disproportionality and the robustness analyzes for the different sub-queries.

**Table S3– Drug-related dysphonia**. Expected signals from the SIDER and unexpected signals from the FAERS were integrated in the following table, together with robustness considerations. Drugs were aggregated based on the ATC classification, Class 1 and Class3. In the FAERS column, robust new signals were reported in **bold**, while non-robust new signals were reported in *italic*.

| ATC level 1 | ATC level 3 | SIDER | FAERS | DAGs & Robustness |
|---|---|---|---|---|
| Nervous | Antidepressants | clomipramine, fluoxetine, citalopram, paroxetine, sertraline, fluvoxamine, escitalopram | | |
| | Antipsychotics | fluphenazine, olanzapine, risperidone, aripiprazole, paliperidone | | |
| | Dopaminergic | ropinirole, pramipexole | | |
| | Anti-Dementia | rivastigmine | | |
| | Parasympathomimetics | pilocarpine, cevimeline | | |
| | Drugs for addictive disorders | nicotine, varenicline, naltrexone | | |
| | Antiepileptics | clonazepam, oxcarbazepine, eslicarbazepine, vigabatrin, topiramate, gabapentin | | |
| | Opioids | nalbuphine | | |
| | Other analgesics | paracetamol | | |
| | Antimigraine | dihydroergotamine, sumatriptan, rizatriptan | | |
| | Anesthetics, general | sevoflurane, fentanyl, remifentanil | | |
| | Anesthetics, local | lidocaine | | |
| | Anxiolytics | alprazolam, buspirone | | |
| | Hypnotics and sedatives | midazolam | | |
| | Other nervous | riluzole | | |
| Cardiovascular | Potassium sparing agents | | **triamterene** | D: ACEI |
| | ACE-Inhibitors | enalapril, perindopril, cilazapril, fosinopril | | |
| | Selective calcium channel blockers, vascular | nifedipine, lercanidipine | | |
| | Beta blockers | bisoprolol | | |
| | Antiarrhythmics I and III | flecainide | | |
| | Selective calcium channel blockers, cardiac | | **diltiazem** | D: ACEI |
| | Lipid modifying agents | | **evolocumab** | D: ACEI |
| Alimentary and Metabolism | Belladonna | atropine, hyoscyamine | | |
| | Propulsives | metoclopramide | | |
| | Anabolic steroids | nandrolone | | |
| | Drugs for peptic ulcer and reflux | lansoprazole | *omeprazole, esomeprazole* | B-C: reflux laryngitis,non fixable |
| | Insulins | insulin detemir | | |
| | Antiemetics and antinauseants | ondansetron | *palonosetron* | C: emesis-related laryngitis non fixable |
| Blood | Antithrombotic | dipyridamole | | |
| | Irrigating solutions | sorbitol | | |
| Hormones | Anti-parathyroid agents | calcitonin | | |
| | Anterior pituitary hormones | | **corticotropin** | |

| | | | | |
|---|---|---|---|---|
| | Hypothalamic hormones | | **octreotide** | |
| Antiinfectives | Quinolone | ofloxacin, ciprofloxacin, lomefloxacin, levofloxacin, trovafloxacin | | |
| | Other Beta-lactam | ertapenem | | |
| | Macrolides | clindamycin | | |
| | Aminoglycoside | tobramycin | **amikacin** | B: infective laryngitis; plausibly not completely fixed because of reports with specified agent but not site of infection |
| | Drugs for tuberculosis | rifapentine | | |
| | Antimycotics, systemic | amphotericin b, itraconazole, posaconazole, caspofungin | | |
| | Direct acting antivirals | foscarnet, zidovudine, zalcitabine, lamivudine, raltegravir, ribavirin, boceprevir | | |
| Respiratory | Decongestans, topical | ipratropium, beclometasone, flunisolide, budesonide, fluticasone, mometasone, ciclesonide, nedocromil, cromoglicic acid | | |
| | Other drugs for obstructive diseases, inhalant | tiotropium, aclidinium | **umeclidinium** | |
| | Antihistamines, systemic | cetirizine, levocetirizine | **rupatadine**, *fexofenadine* | B: allergic respiratory conditions (restricted to urticaria) |
| | Adrenergics, inhalant | salbutamol, pirbuterol, salmeterol, formoterol | **procaterol, indacaterol, olodaterol** | |
| | Other drugs for obstructive diseases | | *theophylline, bamifylline*, **zafirlukast, omalizumab, mepolizumab, benralizumab** | D: inhalants (and C: obstructive conditions) |
| | Expectorants | acetylcysteine, mannitol | *guaifenesin, dornase alfa* | B: cough non fixable |
| Immunomodulating | Hormone antagonists | exemestane | | |
| | Hormones | medroxyprogesterone | | |
| | Protein kinase inhibitors | axitinib, sorafenib, pazopanib, regorafenib, cabozantinib, nilotinib, ponatinib | **lenvatinib, nintedanib, everolimus** | |
| | Monoclonal antibodies | | **bevacizumab, pembrolizumab** | |
| | Antimetabolites | capecitabine | | |
| | Plant alkaloids | vinflunine, paclitaxel | | |
| | Cytotoxic antibiotics | doxorubicin, epirubicin, ixabepilone | | |
| | Other antineoplastic | oxaliplatin, procarbazine, estramustine, bortezomib, celecoxib | **niraparib** | |
| | Immunosuppressants | lenalidomide | **adalimumab** | |
| | Immunostimulants | interferon alfa-2b | | |
| Musculo-skeletal | Drugs affecting bone | ibandronic acid | | |
| | Muscle relaxants, peripherally | | **botulinum toxin** | B: spasmodic dysphonia |
| | Antiinflammatory, non steroids | ibuprofen | | |
| | Muscle relaxants, directly | dantrolene | | |
| Sensory | Antiinfectives | interferon | | |
| | Antiglaucoma | travoprost | | |

| | | | |
|---|---|---|---|
| Dermatologicals | Antipsoriatic, systemic | acitretin | |
| | Anti-acne, topical | isotretinoin | |
| Genitourinary | Urologicals | oxybutynin, solifenacin, trospium, sildenafil | |
| | Other gynecologicals | | **fenoterol** |
| | Androgens | testosterone | |
| | Other sex hormones | danazol | |
| Various | All other therapeutics | protamine, flumazenil | |
| | Other diagnostics | edrophonium | |
| | X-ray contrast media, iodinated | iopromide | |
| | Magnetic resonance imaging contrast media | gadoversetamide | |

**Table S4– Drug-related speech motor control disorder**. Expected signals from the SIDER and unexpected signals from the FAERS were integrated in the following table, together with robustness considerations. Drugs were aggregated based on the ATC classification, Class 1 and Class3. In the FAERS column, robust new signals were reported in **bold**, while non-robust new signals were reported in *italic*.

| ATC level 1 | ATC level 3 | SIDER | FAERS | DAGs & Robustness |
|---|---|---|---|---|
| Nervous | Antipsychotics | perphenazine, ziprasidone, lurasidone, loxapine, clozapine, olanzapine, quetiapine, asenapine, risperidone, aripiprazole, paliperidone, lithium | **haloperidol** | |
| | Anxiolytics | diazepam, oxazepam, lorazepam, clobazam, alprazolam, meprobamate, buspirone | | |
| | Hypnotics and sedatives | flurazepam, nitrazepam, flunitrazepam, triazolam, temazepam, midazolam, zolpidem, zaleplon | | |
| | Antiepileptics | phenytoin, mephenytoin, fosphenytoin, clonazepam, carbamazepine, eslicarbazepine, valproic acid, vigabatrin, tiagabine, lamotrigine, felbamate, topiramate, gabapentin, zonisamide, pregabalin, lacosamide, retigabine, perampanel | | |
| | Anesthetics, general | fentanyl | | |
| | Anesthetics, local | bupivacaine, lidocaine, prilocaine, ropivacaine | | |
| | Opioids | morphine, hydromorphone, buprenorphine, butorphanol, tramadol, tapentadol | | |
| | Antidepressants | amitriptyline, maprotiline, fluoxetine, citalopram, paroxetine, fluvoxamine, escitalopram, moclobemide, trazodone, nefazodone, mirtazapine, bupropion, duloxetine | | |
| | Dopaminergic | amantadine, pramipexole, rasagiline | | |
| | Anticholinergic | | *benzatropine* | B: parkinsonism non fixable |
| | Other analgesics | ziconotide | | |
| | Antimigraine | sumatriptan, rizatriptan | | |
| Cardiovascular | Beta blockers | nadolol | **bisoprolol** | |
| | Thiazides | | **hydrochlorothiazide** | |
| | Selective calcium channel blockers, vascular | | **amlodipine, manidipine, benidipine** | |
| | ACE-Inhibitors | | **ramipril** | |
| | Propulsives | | **metoclopramide** | |

| | | | | |
|---|---|---|---|---|
| Alimentary and Metabolism | Belladonna | atropine, hyoscyamine | | |
| Blood | Antithrombotic | | **clopidogrel, ticlopidine, cilostazol, alteplase** | B: ischaemic stroke-related aphasia |
| Hormones | Corticosteroids, systemic | | **dexamethasone, methylprednisolone** | |
| Antiinfectives | Quinolone | ofloxacin, norfloxacin, levofloxacin | **ciprofloxacin** | |
| | Other antibacterials | metronidazole | | |
| | Direct acting antivirals | aciclovir, valaciclovir, saquinavir | **vidarabine, cidofovir** | |
| Respiratory | Antihistamines, systemic | | **diphenhydramine** | |
| Immunomodulating | Alkylating agents | ifosfamide, lomustine | **cyclophosphamide** | |
| | Antimetabolites | methotrexate, nelarabine, fluorouracil, capecitabine | **mercaptopurine, tioguanine, cytarabine** | |
| | Plant alkaloids | irinotecan | **vincristine** | |
| | Cytotoxic antibiotics | doxorubicin, epirubicin | | |
| | Other antineoplastic | cisplatin, oxaliplatin, procarbazine, bortezomib, pentostatin, mitotane, anagrelide | **asparaginase, pegaspargase** | |
| | Monoclonal antibodies | | **blinatumomab** | |
| | Immunosuppressants | tacrolimus, lenalidomide | *natalizumab,* **eculizumab,** *fingolimod* | C: natalizumab and fingolimod are used in multiple sclerosis, non fixable |
| | Immunostimulants | | *interferon beta-1a* | C: multiple sclerosis non fixable |
| | Hormones | medroxyprogesterone | | |
| Musculo-skeletal | Muscle relaxants, centrally | baclofen, cyclobenzaprine | *tizanidine* | C: multiple sclerosis non fixable |
| | Antiinflammatory, non steroids | indometacin, ibuprofen | **rofecoxib** | |
| Genitourinary | Estrogens | estradiol | | |
| | Progestogens | progesterone | | |

**Table S5– Drug-related aphasia**. Expected signals from the SIDER and unexpected signals from the FAERS were integrated in the following table, together with robustness considerations. Drugs were aggregated based on the ATC classification, Class 1 and Class3. In the FAERS column, robust new signals were reported in **bold**, while non-robust new signals were reported in *italic*.

| ATC level 1 | ATC level 3 | SIDER | FAERS | DAGs & Robustness |
|---|---|---|---|---|
| Nervous | Antipsychotics | clozapine, quetiapine, risperidone, aripiprazole | **lithium** | |
| | Anti-Dementia | donepezil, rivastigmine, galantamine, memantine | | |
| | Dopaminergic | ropinirole, pramipexole, selegiline, rasagiline | | |
| | Anticholinergic | | *procyclidine* | C: Parkinson, non fixable |
| | Antimigraine | sumatriptan, naratriptan, eletriptan | | |
| | Hypnotics and sedatives | zolpidem | | |
| | Anxiolytics | | **diazepam, lorazepam** | |
| | Antiepileptics | lamotrigine, topiramate, gabapentin, pregabalin, retigabine, oxcarbazepine, eslicarbazepine, phenytoin, fosphenytoin | **clonazepam** | |

| | | | | |
|---|---|---|---|---|
| | Antidepressants | clomipramine, citalopram, paroxetine, sertraline, escitalopram, trazodone, mirtazapine, bupropion, venlafaxine, reboxetine | **amitriptyline** | |
| | Anesthetics, general | fentanyl | | |
| | Other analgesics | ziconotide | | |
| | Parasympathomimetics | pilocarpine, cevimeline | | |
| | Drugs for addictive disorders | nicotine | | |
| | Opioids | | **tramadol** | |
| Cardiovascular | Antiarrhythmics I and III | quinidine | | |
| | Beta blockers | bisoprolol | **labetalol** | |
| | Selective calcium channel blockers, vascular | | **nitrendipine** | |
| | Lipid modifying agents | | **atorvastatin** | |
| Alimentary and Metabolism | Propulsives | | **domperidone** | |
| | Blood glucose lowering drugs | gliclazide, glimepiride | | |
| | Drugs for peptic ulcer and reflux | | **omeprazole** | |
| | Antiemetics and antinauseants | | **ondansetron** | |
| Blood | Antithrombotic | | **phenprocoumon, ticlopidine, alteplase** | B: ischaemic stroke-related aphasia |
| Hormones | Corticosteroids, systemic | | **methylprednisolone** | |
| | Thyroid preparations | | **levothyroxine** | |
| Antiinfectives | Quinolone | ofloxacin, ciprofloxacin, norfloxacin, lomefloxacin, sparfloxacin, levofloxacin, moxifloxacin, gemifloxacin | | |
| | Antimycotics, systemic | posaconazole | | |
| | Drugs for tuberculosis | rifabutin | | |
| | Direct acting antivirals | ganciclovir, foscarnet, ritonavir, zalcitabine | | |
| | Other Beta-lactam | | **cefepime** | |
| Immunomodulating | Immunosuppressants | lenalidomide, tacrolimus | *fingolimod, natalizumab* | C: mulitple sclerosis, non fixable |
| | Immunostimulants | glatiramer, interferon alfa-2b | *interferon beta-1a, peginterferon beta-1a* | C: mulitple sclerosis, non fixable |
| | Alkylating agents | carmustine, temozolomide | **cyclophosphamide, ifosfamide** | C: brain tumor |
| | Antimetabolites | methotrexate, nelarabine, capecitabine | **fludarabine, cytarabine, fluorouracil** | C: brain tumor |
| | Other antineoplastic | oxaliplatin | **pegaspargase, axicabtagene ciloleucel, tisagenlecleucel-t** | C: brain tumor |
| | Plant alkaloids | | **vincristine, etoposide, irinotecan** | C: brain tumor |
| | Cytotoxic antibiotics | | **daunorubicin, mitoxantrone** | C: brain tumor |
| | Protein kinase inhibitors | | **erlotinib, dabrafenib, trametinib, avapritinib** | C: brain tumor |
| | Monoclonal antibodies | | **bevacizumab, blinatumomab** | C: brain tumor |
| Musculo-skeletal | Antiinflammatory, non steroids | diclofenac | | |
| Sensory | Diagnostic agents | fluorescein | | |

| | | | |
|---|---|---|---|
| | Antiglaucoma | | **diclofenamide** |
| Dermatologicals | Anti-acne, topical | isotretinoin | |
| Genitourinary | Estrogens | estradiol | |
| | Antiandrogens | cyproterone | |
| | Hormonal contraceptives | | **drospirenone** |
| Various | X-ray contrast media, iodinated | iopamidol, iopromide, ioversol | |
| | Magnetic resonance imaging contrast media | gadobutrol | |
| | All other therapeutics | deferoxamine | |
| Antiparasitic | Antimalarials | quinine | **mefloquine** |
| | Antinematodals | | **ivermectin** |

**Table S6– Drug-related stereotypies**. Expected signals from the SIDER and unexpected signals from the FAERS were integrated in the following table, together with robustness considerations. Drugs were aggregated based on the ATC classification, Class 1 and Class3. In the FAERS column, robust new signals were reported in **bold**, while non-robust new signals were reported in *italic*.

| ATC level 1 | ATC level 3 | SIDER | FAERS | DAGs & Robustness |
|---|---|---|---|---|
| Nervous | Antiepileptics | topiramate | | |
| | Antidepressants | phenelzine, bupropion | | |
| Immunomodulating | Alkylating agents | ifosfamide | | |
| Various | X-ray contrast media, iodinated | iopamidol | | |

**Table S7– Drug-related tachyphrenia**. Expected signals from the SIDER and unexpected signals from the FAERS were integrated in the following table, together with robustness considerations. Drugs were aggregated based on the ATC classification, Class 1 and Class3. In the FAERS column, robust new signals were reported in **bold**, while non-robust new signals were reported in *italic*.

| ATC level 1 | ATC level 3 | SIDER | FAERS | DAGs & Robustness |
|---|---|---|---|---|
| Nervous | Antiepileptics | carbamazepine | **valproic acid, lamotrigine** | |
| | Dopaminergic | pramipexole | | |
| | Antipsychotics | aripiprazole | **lurasidone, quetiapine, lithium** | |
| | Anxiolytics | alprazolam | | |
| | Hypnotics and sedatives | flurazepam | | |
| | Antidepressants | duloxetine | | |
| | Psychostimulants | methylphenidate, dexmethylphenidate, lisdexamfetamine | | |
| Hormones | Thyroid preparations | | **levothyroxine** | |
| Immunomodulating | Alkylating agents | ifosfamide | | |
| | Other antineoplastic | | **niraparib** | |
| Musculo-skeletal | Antiinflammatory, non steroids | | **ibuprofen** | |
| Antiparasitic | Antinematodals | | **ivermectin** | |

**Table S8– Drug-related bradyphrenia**. Expected signals from the SIDER and unexpected signals from the FAERS were integrated in the following table, together with robustness considerations. Drugs were aggregated based on the ATC classification, Class 1 and Class3. In the FAERS column, robust new signals were reported in **bold**, while non-robust new signals were reported in *italic*.

| ATC level 1 | ATC level 3 | SIDER | FAERS | DAGs & Robustness |
|---|---|---|---|---|
| Nervous | Other analgesics | ziconotide | | |
| | Antimigraine | clonidine | | |
| | Antiepileptics | topiramate, zonisamide, pregabalin | **clonazepam, valproic acid** | |
| | Dopaminergic | pramipexole | | |
| | Antipsychotics | ziprasidone, aripiprazole | **olanzapine, lithium** | |
| | Anxiolytics | alprazolam | | |
| | Hypnotics and sedatives | | **lormetazepam** | |
| Alimentary and Metabolism | Antiobesity | | **lorcaserin** | |
| Immunomodulating | Alkylating agents | ifosfamide | | |
| Genitourinary | Estrogens | estradiol | | |

**Table S9– Drug-related incoherence**. Expected signals from the SIDER and unexpected signals from the FAERS were integrated in the following table, together with robustness considerations. Drugs were aggregated based on the ATC classification, Class 1 and Class3. In the FAERS column, robust new signals were reported in **bold**, while non-robust new signals were reported in *italic*.

| ATC level 1 | ATC level 3 | SIDER | FAERS | DAGs & Robustness |
|---|---|---|---|---|
| Nervous | Anxiolytics | diazepam, lorazepam, buspirone | | |
| | Hypnotics and sedatives | amobarbital, secobarbital, flurazepam, nitrazepam, estazolam, triazolam, quazepam, zopiclone, zolpidem, zaleplon, eszopiclone, ramelteon, suvorexant | | |
| | Antidepressants | clomipramine, doxepin, fluoxetine, citalopram, paroxetine, sertraline, fluvoxamine, escitalopram, nefazodone, mirtazapine, bupropion, venlafaxine, reboxetine | | |
| | Psychostimulants | methylphenidate, modafinil, dexmethylphenidate | | |
| | Parasympathomimetics | pilocarpine, cevimeline | | |
| | Drugs for addictive disorders | varenicline, acamprosate, naltrexone | | |
| | Dopaminergic | pergolide, ropinirole, selegiline, tolcapone, entacapone | | |
| | Antipsychotics | olanzapine, quetiapine, risperidone, aripiprazole | | |
| | Antiepileptics | phenobarbital, phenytoin, fosphenytoin, carbamazepine, oxcarbazepine, valproic acid, vigabatrin, tiagabine, felbamate, topiramate, gabapentin, levetiracetam, pregabalin | | |
| | Anesthetics, general | fentanyl, propofol, oxybate sodium | | |
| | Opioids | morphine, oxycodone, butorphanol, tramadol, tapentadol | | |
| | Antimigraine | zolmitriptan, eletriptan, frovatriptan | | |
| | Anti-Dementia | memantine | | |
| | Other nervous | riluzole | | |
| Cardiovascular | Cardiac stimulants | | **droxidopa** | |
| | Beta blockers | betaxolol, esmolol, carvedilol | | |
| | Antiadrenergic, peripherically acting | doxazosin | | |
| | Selective calcium channel blockers, cardiac | diltiazem | | |

| | | | |
|---|---|---|---|
| | Selective calcium channel blockers, vascular | nisoldipine | |
| Alimentary and Metabolism | Drugs for peptic ulcer and reflux | pantoprazole | |
| | Antiemetics and antinauseants | dronabinol, nabilone | |
| | Antiobesity | sibutramine | |
| Blood | Antithrombotic | | **ticagrelor** |
| Hormones | Posterior pituitary hormones | desmopressin | |
| | Thyroid preparations | | **levothyroxine, liothyronine** |
| Antiinfectives | Quinolone | lomefloxacin, sparfloxacin, trovafloxacin, moxifloxacin, gatifloxacin | |
| | Antimycotics, systemic | amphotericin b | |
| | Direct acting antivirals | ganciclovir, valganciclovir, ritonavir, stavudine, efavirenz | **zanamivir** |
| Respiratory | Antihistamines, systemic | cetirizine, levocetirizine | **diphenhydramine** |
| | Decongestans, topical | azelastine | |
| | Adrenergics, inhalant | salbutamol | |
| | Expectorants | acetylcysteine | |
| Immunomodulating | Alkylating agents | carmustine | |
| | Plant alkaloids | paclitaxel | |
| | Cytotoxic antibiotics | doxorubicin, daunorubicin, epirubicin | |
| | Other antineoplastic | bexarotene, pentostatin | |
| | Hormones | megestrol, leuprorelin, goserelin | |
| | Immunostimulants | interferon alfa-2b, glatiramer | **peginterferon alfa-2b** |
| | Immunosuppressants | mycophenolic acid, tacrolimus, thalidomide | **siponimod** |
| Sensory | Mydriatics | cyclopentolate, homatropine | |
| Dermatologicals | Antibiotics, topical | | **rifaximin** |
| Genitourinary | Urologicals | oxybutynin, darifenacin | |
| Antiparasitic | Antimalarials | | **mefloquine** |
| | Antinematodals | | **ivermectin** |

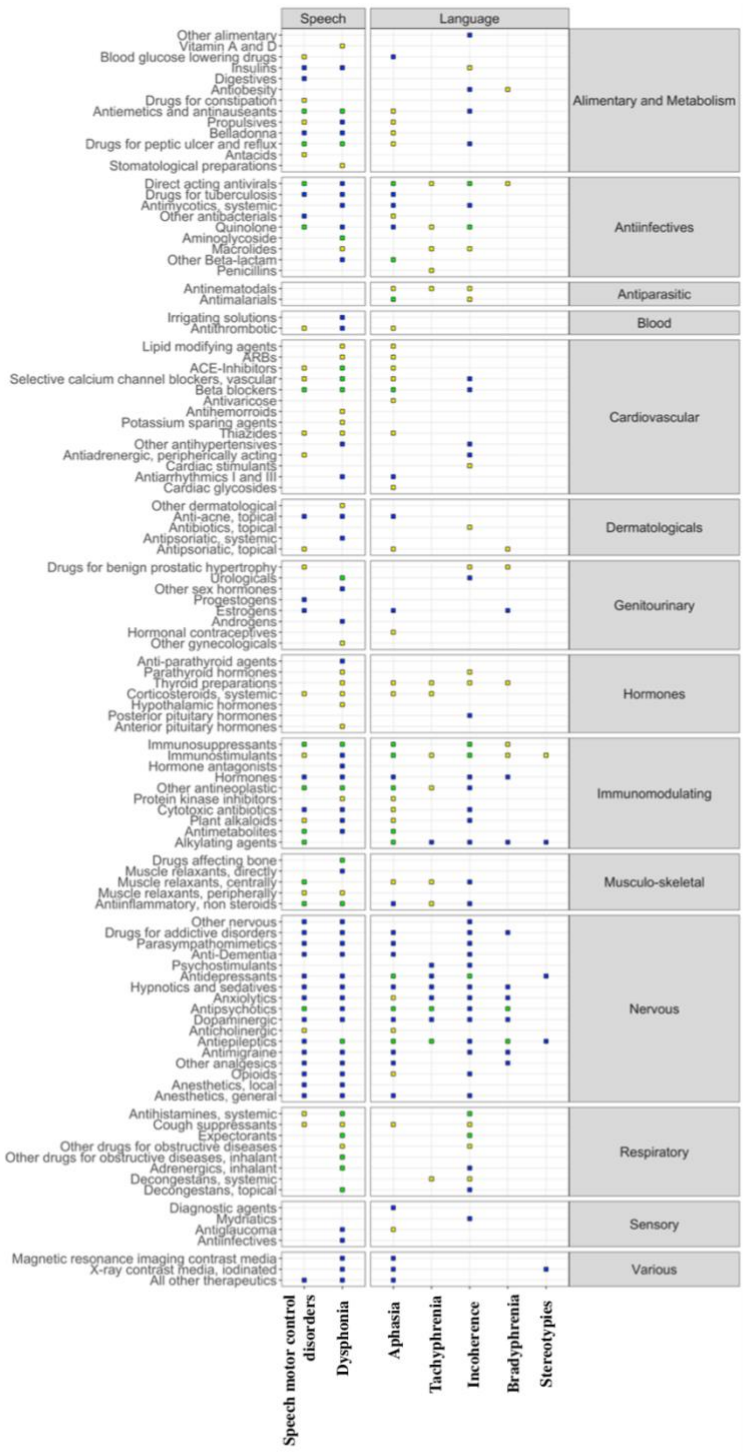

**Figure S6 – Heat Map of drug-related communicative atypicalities.** We showed expected and unexpected associations between drug classes at the third level of the ATC classification (y-axis – left, aggregated by the first level of the ATC classification – right) and communicative impairment sub-queries (x-axis – bottom, aggregated in Speech and Language – top). Blue squares represent expected associations based on SIDER, yellow squares unexpected associations, and green squares drug classes that included both expected and unexpected medications. Rows and columns with no significant association were not shown.

**Table S10 – List of drugs associated with communicative atypicalities.** Expected drugs are those already identified in SIDER. Unexpected drugs are split in 2 classes: new or integrated, for which drugs within the same class are already known to associate with the impairment. *Robust FAERS signals*.

| | DYSPHONIA | SPEECH MOTOR CONTROL | APHASIA | STEREOTYPIES | TACHYPHRENIA/ BRADYPHRENIA | INCOHERENCE |
|---|---|---|---|---|---|---|
| SUMMARY DRUG CLASS | 53 expected, 10 integrated, 9 new | 17 expected, 10 integrated, 10 new | 19 expected, 10 integrated, 22 new | 4 expected | 8 expected, 3 integrated, 5 new | 34 expected, 4 integrated, 6 new |
| **SIDER** | clomipramine, fluoxetine, citalopram, paroxetine, sertraline, fluvoxamine, escitalopram, fluphenazine, olanzapine, risperidone, aripiprazole, paliperidone, ropinirole, pramipexole, rivastigmine, pilocarpine, cevimeline,nicotine, varenicline, naltrexone, clonazepam, oxcarbazepine, eslicarbazepine, vigabatrin, topiramate, gabapentin, nalbuphine, paracetamol, dihydroergotamine, sumatriptan, rizatriptan, sevoflurane, fentanyl, remifentanil, lidocaine, alprazolam, buspirone, midazolam, riluzole, enalapril, perindopril, cilazapril, fosinopril, nifedipine, lercanidipine, bisoprolol, flecainide, atropine, hyoscyamine, metoclopramide, nandrolone, lansoprazole, insulin detemir, ondansetron, dipyridamole, sorbitol, calcitonin, ofloxacin, ciprofloxacin, lomefloxacin, levofloxacin, trovafloxacin, ertapenem, clindamycin, tobramycin, rifapentine, amphotericin b, itraconazole, posaconazole, caspofungin, foscarnet, zidovudine, zalcitabine, lamivudine, raltegravir, ribavirin, boceprevir, ipratropium, beclometasone, flunisolide, budesonide, fluticasone, mometasone, ciclesonide, nedocromil, cromoglicic acid, tiotropium, aclidinium, cetirizine, levocetirizine, salbutamol, pirbuterol, salmeterol, formoterol, acetylcysteine, mannitol, exemestane, medroxyprogesterone, axitinib, sorafenib, pazopanib, | perphenazine, ziprasidone, lurasidone, loxapine, clozapine, olanzapine, quetiapine, asenapine, risperidone, aripiprazole, paliperidone, lithium, diazepam, oxazepam, lorazepam, clobazam, alprazolam, meprobamate, buspirone, flurazepam, nitrazepam, flunitrazepam, triazolam, temazepam, midazolam, zolpidem, zaleplon, phenytoin, mephenytoin, fosphenytoin, clonazepam, carbamazepine, eslicarbazepine, valproic acid, vigabatrin, tiagabine, lamotrigine, felbamate, topiramate, gabapentin, zonisamide, pregabalin, lacosamide, retigabine, perampanel, fentanyl, bupivacaine, lidocaine, prilocaine, ropivacaine, morphine, hydromorphone, buprenorphine, butorphanol, tramadol, tapentadol, amitriptyline, maprotiline, fluoxetine, citalopram, paroxetine, fluvoxamine, escitalopram, moclobemide, trazodone, nefazodone, mirtazapine, bupropion, duloxetine, amantadine, pramipexole, rasagiline, ziconotide, sumatriptan, rizatriptan, nadolol | clozapine, quetiapine, risperidone, aripiprazole,donepezil, rivastigmine, galantamine, memantine,ropinirole, pramipexole, selegiline, rasagiline, sumatriptan, naratriptan, eletriptan,zolpidem, lamotrigine, topiramate, gabapentin, pregabalin, retigabine, oxcarbazepine, eslicarbazepine, phenytoin, fosphenytoin,clomipramine, citalopram, paroxetine, sertraline, escitalopram, trazodone, mirtazapine, bupropion, venlafaxine, reboxetine,fentanyl,ziconotide, pilocarpine, cevimeline, nicotine, quinidine, bisoprolol, gliclazide, glimepiride, ofloxacin, ciprofloxacin, norfloxacin, lomefloxacin, sparfloxacin, levofloxacin, moxifloxacin, gemifloxacin, posaconazole, rifabutin, ganciclovir, foscarnet, ritonavir, zalcitabine, lenalidomide, tacrolimus, glatiramer, interferon alfa-2b, carmustine, temozolomide<br>methotrexate, nelarabine, capecitabine, oxaliplatin, diclofenac, fluorescein, isotretinoin, estradiol, cyproterone, iopamidol, iopromide, ioversol, gadobutrol, deferoxamine, quinine | topiramate, phenelzine, bupropion, ifosfamide, iopamidol | ziconotide, clonidine, carbamazepine, topiramate, zonisamide, pregabalin, pramipexole, ziprasidone, aripiprazole, alprazolam, flurazepam, duloxetine, methylphenidate, dexmethylphenidate, lisdexamfetamine, ifosfamide, estradiol | diazepam, lorazepam, buspirone, amobarbital, secobarbital, flurazepam, nitrazepam, estazolam, triazolam, quazepam, zopiclone, zolpidem, zaleplon, eszopiclone, ramelteon, suvorexant, clomipramine, doxepin, fluoxetine, citalopram, paroxetine, sertraline, fluvoxamine, escitalopram, nefazodone, mirtazapine, bupropion, venlafaxine, reboxetine, methylphenidate, modafinil, dexmethylphenidate, pilocarpine, cevimeline, varenicline, acamprosate, naltrexone, pergolide, ropinirole, selegiline, tolcapone, entacapone, olanzapine, quetiapine, risperidone, aripiprazole, phenobarbital, phenytoin, fosphenytoin, carbamazepine, oxcarbazepine, valproic acid, vigabatrin, tiagabine, felbamate, topiramate, gabapentin, levetiracetam, pregabalin, fentanyl, propofol, oxybate sodium, morphine, oxycodone, butorphanol, tramadol, tapentadol, zolmitriptan, eletriptan, frovatriptan, memantine, riluzole, betaxolol, esmolol, carvedilol, doxazosin, diltiazem, nisoldipine, pantoprazole, dronabinol, nabilone, sibutramine, desmopressin, lomefloxacin, sparfloxacin, trovafloxacin, moxifloxacin, gatifloxacin, amphotericin b, ganciclovir, valganciclovir, ritonavir, stavudine, efavirenz, cetirizine, levocetirizine, azelastine<br>salbutamol, acetylcysteine, carmustine, paclitaxel, doxorubicin, daunorubicin, |

| | | | | | | |
|---|---|---|---|---|---|---|
| | regorafenib, cabozantinib, nilotinib, ponatinib, capecitabine, vinflunine, paclitaxel, doxorubicin, epirubicin, ixabepilone, oxaliplatin, procarbazine, estramustine, bortezomib, celecoxib, lenalidomide, interferon alfa-2b, ibandronic acid, ibuprofen, dantrolene, interferon, travoprost, acitretin, isotretinoin, oxybutynin, solifenacin, trospium, sildenafil, testosterone, danazol, protamine, flumazenil, edrophonium, iopromide, gadoversetamide | | | | | epirubicin, bexarotene, pentostatin, megestrol, leuprorelin, goserelin, interferon alfa-2b, glatiramer, mycophenolic acid, tacrolimus, thalidomide, cyclopentolate, homatropine, oxybutynin, darifenacin |
| **FAERS** | * triamterene, diltiazem, evolocumab, corticotropin, octreotide, amikacin, umeclidinium, rupatadine, procaterol, indacaterol, olodaterol, zafirlukast, omalizumab, mepolizumab, benralizumab, lenvatinib, nintedanib, everolimus, bevacizumab, pembrolizumab, niraparib, adalimumab, botulinum toxin, fenoterol *<br>omeprazole, esomeprazole, palonosetron, fexofenadine, theophylline, bamifylline, guaifenesin, dornase alfa | * haloperidol, benzatropine, bisoprolol, hydrochlorothiazide, amlodipine, manidipine, benidipine, ramipril, metoclopramide, clopidogrel, ticlopidine, cilostazol, alteplase, dexamethasone, methylprednisolone, ciprofloxacin, vidarabine, cidofovir, diphenhydramine, cyclophosphamide, mercaptopurine, tioguanine, cytarabine, vincristine, asparaginase, pegaspargase, blinatumomab, eculizumab, rofecoxib *<br><br>natalizumab, fingolimod, interferon beta-1a,<br>tizanidine | * lithium, diazepam, lorazepam, clonazepam, amitriptyline, tramadol, labetalol, nitrendipine, atorvastatin, domperidone, omeprazole, ondansetron, phenprocoumon, ticlopidine, alteplase, methylprednisolone, levothyroxine, cefepime, cyclophosphamide, ifosfamide, fludarabine, cytarabine, fluorouracil, pegaspargase, axicabtagene ciloleucel, tisagenlecleucel-t, vincristine, etoposide, irinotecan, daunorubicin, mitoxantrone, erlotinib, dabrafenib, trametinib, avapritinib, bevacizumab, blinatumomab, diclofenamide, drospirenone, mefloquine, ivermectin *<br><br>procyclidine, fingolimod, natalizumab, interferon, beta-1a, peginterferon beta-1a | | * valproic acid, lamotrigine, clonazepam, lurasidone, quetiapine, olanzapine, lithium, lormetazepam, levothyroxine, niraparib, ibuprofen, ivermectin, lorcaserin * | * droxidopa, ticagrelor, levothyroxine, liothyronine, zanamivir, diphenhydramine, peginterferon alfa-2b, siponimod, rifaximin, mefloquine, ivermectin * |
| **PROPOSED MECHANISM** | Anticholinergic toxicity, Extrapyramidal dystonia, laryngeal irritation, vocal cord thickening, anti-angiogenetic action, cytotoxicity, vocal cord hemorrages, vocal cord nodules | Extrapyramidal dystonia, sedation, neurotoxicity, stroke, dopaminergic, catecholaminergic and GABAergic pathways | Neurotoxicity, Blood brain barrier permeability, dopamine antagonism, stroke | | Altered arousal | |
